# Towards reproducible multimorbidity clustering in electronic health records: a transparent pipeline for aligning research aims and methodology

**DOI:** 10.64898/2026.05.25.26353178

**Authors:** Guillermo Romero Moreno, Valerio Restocchi, Luna De Ferrari, Jake Palmer, Jacques Fleuriot, Bruce Guthrie, Nazir Lone

**Author notes:** Equal contribution (Bruce Guthrie, Nazir Lone).

## Abstract

The availability of electronic health records has facilitated data-driven approaches to the understanding of multimorbidity, with clustering becoming a common tool for uncovering relevant groups of associated conditions. Previous studies, however, have found challenges in their reproducibility, with wide disparity in the reported clusters. At the core of this issue lays a vagueness of the definition of a cluster, leading to a lack of standards in their methods and evaluation, while implementation details are often not completely reported or explicit in their assumptions. We present a methodological pipeline that can be adapted to different cluster definitions (e.g. multiple cluster membership or clusters where all nodes are mutually associated) and a set of scores that can be composed into an evaluation metric that explicitly incorporates assumptions that align with the research aims. We apply our pipeline to a healthcare dataset of over 7 million patients in England and show how clusters may drastically differ when varying the parameter choices, exposing the risks of reporting a single clustering realisation. Our methodological pipeline, evaluation framework, and tools for analysis and network visualisation serve as a reference to transparently explore and align methodological decisions to the aims of multimorbidity clustering, contributing to overcome the reproducibility challenges of the field.

## Introduction

The prevalence of multimorbidity, defined as the co-occurrence of two or more long-term conditions, is increasing globally [1]. People with multimorbidity often require greater support from health and social care systems, placing greater pressures on these services [2]. Defining multimorbidity by counting the number of long-term conditions has the advantage of being simple to operationalise and easily understandable [3]. However, a more complex and nuanced approach is required to facilitate a deeper understanding of the causes of multimorbidity and of its impact on individuals and health systems. Recently, approaches relying on big data have been growingly applied to the understanding of multimorbidity [4–6], such as grouping long-term conditions into clusters [7] or examining how conditions relate to each other from networks of association [8–10].

Identifying clusters of long-term conditions presents its own challenges. For example, the theoretical number of unique groups of two, three or four conditions drawn from the Elixhauser list [B] of 30 long-term conditions is 31,898 and from the Delphi consensus list [12] of 59 long-term conditions is 489,346. Therefore, one of the aims of clustering of long-term conditions is to translate the inherent complexity of between-condition associations into clinically relevant solutions for identifying common aetiological mechanism or for use in outcome prediction [13,14].

However, the use of clustering for the study of multimorbidity has been characterised by a wide disparity of findings with important challenges in reproducibility [4,5,15]. Although some of these challenges stem from the complex management of restricted Electronic Health Records [16], equal barriers stem from inconsistencies and lack of standards in the methodological approaches used for clustering [4,6,15]. Regardless of the approach chosen, the methodological pipeline for developing networks and finding clusters requires multiple steps, where decisions taken at each of them may be critical in shaping how the resulting clusters look like [17–19].

These decisions are not only often incompletely reported, but very rarely supported by an explicit theoretical rationale that links them to the research aims of the clustering [4,20,21]. For instance, clustering solutions should account for chance associations within a network so that only non-random clusters are identified [8,10,22,23]. Other features of clustering solutions that should be considered include the number and size of clusters, whether to allow conditions to belong to more than one cluster, and whether all conditions within a cluster must be directly associated with each other [15,23].

Furthermore, multimorbidity clustering studies often show only one realisation of the results using a single set of parameters without acknowledging how different parameter choices might yield alternative results [17,19]. Last, multimorbidity clustering research suffers from a lack of agreed-upon approaches in how to evaluate solutions [6,24], stemming from a lack of a gold standard and, more fundamentally, a common understanding of what a cluster even means [15,20].

Here, we present a methodological pipeline for finding non-random clusters of conditions from electronic health records, where conditions may have multiple cluster memberships and with the possibility of requiring all conditions within a cluster to be mutually associated. We apply it to a primary care dataset of over 7 million patients in England and demonstrate how varying parameters in the methodology and decisions on cluster definitions affects results, while also offering alternative visualisations that facilitate clinical inspection.

To facilitate evaluation, we propose a set of scores that characterises different aspects of the clustering solutions and a systematic approach for selecting one solution via a combination of these scores. This criterion not only is reproducible and interpretable but can also be easily tailored to the definition of multimorbidity clustering that best aligns with the research aims [15]. Last, we expose some of the risks in only reporting a single ‘best’ solution [15,25], as very different and nearly-as-good solutions may coexist for different parameter combinations, a problem that grows substantially with the number of parameters in the pipeline.

## Methods

### Study population

We performed a cross-sectional study on an electronic-health-records representative dataset for our study, the Clinical Practice Research Datalink (CPRD) Aurum, which includes primary care data as well as linked data for more than 40 million patients in England [26]. This data was additionally enriched with a linked hospital dataset, Hospital Episode Statistics (HES). Aside from medical and administrative data, the CPRD Aurum dataset includes basic demographic information, such as age, sex, ethnicity, and socioeconomic deprivation.

Eligible participants were those aged between 30 years and 99 years on the 1st of January 2018, who had been permanently registered with an up to standard primary care practice, and with up to standard individual data for at least one year (to account for falsely inflated incidence at or soon after registration from date errors [27]). Due to their small numbers and unreliability in recording, we removed individuals with ‘Undefined’ or ‘I’ in the sex field (125, <0.002%). This resulted in 7,490,874 individuals in our study population.

We stratified the population by sex and age in ten-year intervals except for the oldest group: 30-39, 40-49, 50-59, 60-69, 70-79, 80-99, where age was taken on the 1^st^ of January of 2018.

More details in the processing steps of the CPRD dataset can be found in supplementary material S10.

### Condition definition

Our study drew on a list of 79 long-term conditions compiled by clinicians in the research team. Only conditions with a recorded date up to 1 January 2018 were considered. The list ensured full coverage of conditions recommended by a recent consensus study for multimorbidity research where routine data permit valid measurement, and was tailored to meet the specific aims and analytical framework of the present work [12]. Condition status was ascertained through coded diagnoses, using Read v2 terminology in primary care data and ICD-10 classifications in hospital datasets, supplemented where appropriate by laboratory tests and measurements. We only allowed the condition ‘Bening prostate hyperplasia’ for men and ‘Endometriosis’ and ‘Polycystic ovary syndrome’ for women, so we removed such diagnoses when present in the opposite sex. This resulted in 77 conditions being retained for men and 78 for women (see supplementary material S1 for the full list of conditions).

### Clustering pipeline

Departing from the cleaned dataset, our clustering approach contains several steps, as can be seen in Figure 1. We first extract pairwise associations between health conditions, then build multimorbidity networks from the association data, and then apply community detection to find clusters of conditions tightly associated.

**Figure 1.**
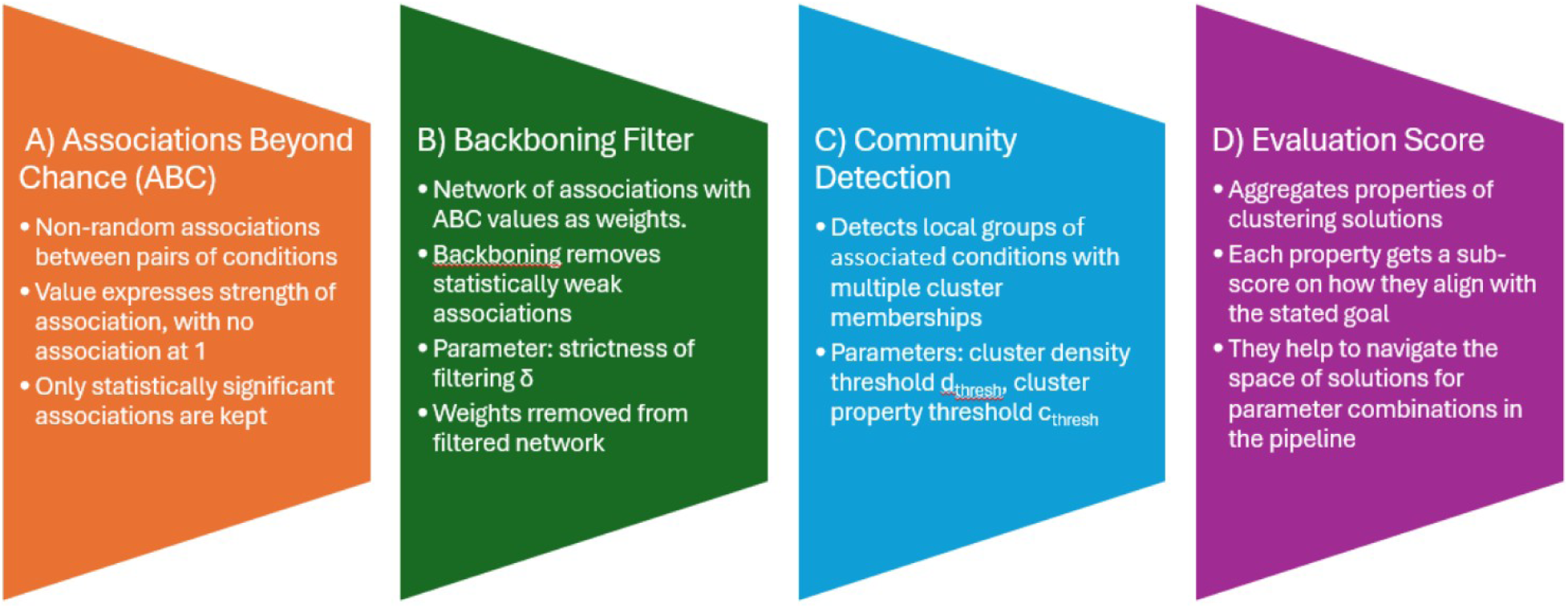
Methodological pipeline for multimorbidity clustering.

To infer associations between health conditions, we used the Association Beyond Chance (ABC) framework [22], which provides Bayesian estimation of associations from count and co-occurrence data while controlling for uncertainty when counts are low. An ABC value between two conditions of ABC=1 determines that their association is no different to what would be expected at random; values ABC>1 determine how many times they are positively associated beyond chance, and ABC<1 imply a mutually exclusive behaviour -- up to a minimum of ABC=0 if the two conditions never co-exist. The ABC framework is Bayesian and outputs a posterior distribution of ABC values per association, in the form of samples. To determine the association value *A_ij_* between a pair of conditions, we use the estimated mode of the distribution of samples, and we determine whether the association is significant if 99% of the samples are above (or below) one [22].

As some conditions are mutually exclusive by definition (i.e., Rheumatoid Arthritis and Connective tissue disorder; Diabetes Type 1 and Type 2), we forbid their association and treat them as non-significant. We select prior distributions for ABC values as log-normal distributions with mean 0.25 and std dev 0.5 (i.e., with their mode at one) and sample 1000 points from the posterior distributions, after 2000 warm-up steps.

We then build multimorbidity networks, where each condition is a node and edges connecting conditions represent whether a significant association exists between them, with a weight corresponding to the estimated *A_ij_* association value. As most associations are significant (even if weak), leading to very dense networks, we apply a Noise-Corrected backboning filter [28] that only preserves connections whose association values are above what is expected by chance from the average association patterns. The backboning filter has a parameter *δ* ∈ *R* that controls the strictness of the filtering. To remove condition prevalence biases that are present in association values [8,29], we discard the weight information (*A_ij_*) and work with the resulting unweighted network. We remove conditions from the network if all their associations have been filtered out.

Last, we apply a community detection algorithm to the filtered network that finds groups of nodes tightly associated. We use DPClus [30], a community detection algorithm that allows for multiple group memberships for the same condition and follows a local approach, i.e., cluster decision only concern nodes and their neighbours and are not affected by distant parts of the network. DPClus starts from ‘seed nodes’ and uses then two cluster-related measures to consider whether neighbouring nodes should be added to a cluster: 1) the density of a cluster *d_k_*, which is defined by the ratio between the number of edges present within the cluster and the theoretically maximum number of edges that could be present, and the 2) *cluster-property* between a node and a cluster *cp_nk_*, which is the ratio between the number of edges joining the node to the cluster and the expected number of edges that would be necessary to keep the density of the cluster unchanged.

For a neighbouring node to enter a cluster, the values of *d_k_* and *cp_nk_* need to be higher than pre-specified thresholds *d_thresh_* and *cp_thresh_*, which are the two main parameters of the algorithm, and which control how strict cluster definitions are. Note that the strictest extreme of *d_thresh_=1* would enforce that all conditions within a cluster must be directly associated (after filtering) with every other condition within the cluster. This would correspond to a ‘closed’ group [23] and reflects the strongest possible cohesiveness within a cluster, potentially reflecting a single factor simultaneously all conditions in the group. Also note that with this clustering approach conditions may not belong to any cluster, if their connection patterns are not strong enough to meet the density and cluster-property criteria. We used the DPClus implementation from the python package CDLib [31] version 0.4.0.

We run two sets of experiments in which we analyse how varying some parameters in the pipeline affects the resulting clustering solutions. In the first set of experiments, we only focus on closed clusters where every condition must be associated with all others within the cluster. The rationale for this approach is to increase internal coherence within each cluster and to identify tightly linked multimorbidity patterns that reflect robust and potentially causal relationships. To implement this, we fix the parameter *d_thresh_=1* and we vary the backboning parameter *δ* from –2 to +6, in steps of 0.02, with a total of 401 parameter settings.

In the second set of experiments, we allow the possibility of ‘open’ clusters and jointly vary the backboning parameter *δ* from –1.5 to +3.5, in steps of 0.05, and the clustering parameter *d_thresh_* from 0 to 1 in steps of 0.01, with a total of 10,201 parameter combinations. We run each parameter combination on every age-sex stratum. For simplicity, we fix the clustering parameter *cp_thresh_* at 0.5 in all the experiments shown in the main text; we show how it affects clustering solution in the supplementary material (Figure S8).

The code implementing the pipeline and to reproduce all results of the paper can be found at https://github.com/Juillermo/MMC-pipeline/.

### Evaluation of clustering solutions

Due to the exploratory nature of clustering, there is no gold standard against which to compare results, which has led to a variety of proxies to assess the quality of results, but with little consensus or consistency in their use [24]. One fundamental reason for this is that there is not even a consensus on what a cluster means, leading to very disparate results due to the different cluster conceptualisation that algorithms implicitly assume [23], while ‘generic performance metrics’ often convey another set of assumptions that are rarely analysed and translated to the problem of multimorbidity clustering [15,20].

To facilitate cluster evaluation within the context of multimorbidity clustering, we propose the *Multimorbidity Clustering Score* (*MCS*), which reflects some assumptions regarding the characteristics of the multimorbidity clusters that we are looking for, and what properties of the resulting networks and clustering are desirable.

Our proposed *MCS* is composed of the following sub-scores:

- Number of conditions in the network (*N*).
- The average number of edges per node (*E*).
- Numbers of clusters (*C*).
- Number of unclustered conditions (*U*).
- Size of the biggest cluster (*S*).
- Number of conditions with multiple memberships (*O*).
- The stability of the solution (*R*).

All the sub-scores are combined into the *MCS* as

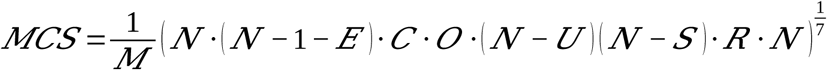

where *M* represents the total number of conditions in the data (77 in men, 78 in women), confining the *MCS* to the interval [0,1]. The multiplicative form of the expression along with the 1/7 exponent encapsule a geometric mean of the seven sub-scores, implying that if any of the criteria goes to its unfavourable extreme, the *MCS* goes down to zero and the solution becomes automatically undesirable.

The stability of the solution (*R*) reflects differences in the resulting clusters when small variations are applied to the methodological parameters, penalising solutions that may arise as artifacts from very specific methodological combinations due to numerical instabilities. We measure the difference between two clustering solutions with the *Overlapping Normalised Mutual Information* (*ONMI*) [32], which falls within the [0,1] range and reaches a value of 1 when the two solutions are identical. We use the *ONMI* to compare a solution with those obtained by varying the parameters (backboning parameter *δ* or clustering parameter *d_thresh_*) in small steps (up to five) and define the stability of the solution (*R*) as the minimum *ONMI* thus obtained. Note that this sub-score is sensitive to the number and size of steps chosen, which in our case correspond to those for exploring the parameter space, as explained above. Other variations in the definition of *R* are shown in the supplementary material (Figures S3 and S9).

The *MCS* allows us to select acceptable solutions across methodological decisions in our pipeline regarding different parameter combinations of the backboning filter and clustering algorithm.

The sub-scores *N* (number of conditions in the network) and *E* (average number of edges per node) are only affected by the backboning step in the pipeline, and they favour networks that are comprehensive (i.e., including as many conditions as possible) but ‘lean’ (fewer edges are preferred).

The sub-scores *C* (number of clusters), *U* (number of unclustered conditions), and *S* (size of the biggest cluster) favour clustering solutions that cover the whole set of conditions with multiple, local clusters, while penalising solutions that include a big, catch-all clusters.

The sub-score *O* (number of conditions with multiple memberships) favours solutions where clusters overlap, as this is a property that we deem desirable from a clinical perspective, given that it is common for conditions to be present in more than one clinical pathway or context.

### Network visualisation

Network visualisations are made using the SFDP spring-block layout algorithm [33], with edge width between nodes *i* and *j* proportional to *|log(ABC_ij_)|* and node size (area) of condition *i* with prevalence *P_i_* proportional to -(*log(P_i_) – min_k_(log(P_k_)))*.

### Code availability

The underlying code and network data for this study are available in GitHub and can be accessed via this link:

https://github.com/Juillermo/MMC-pipeline/tree/main.

### Ethical statement

The use of CPRD Aurum data was approved by the CPRD Research Data Governance Process (Protocol 21_000542). Individual patients can opt out of sharing their data for research and CPRD does not collect data for these patients.

## Results

### Solutions with ‘closed’ clusters

We first present results with ‘closed’ clusters, meaning that all nodes in a cluster have direct associations to every other node within the cluster. With the clustering algorithm from our pipeline, this is achieved when the density threshold is fixed at *dthresh* =1.

For closed clustering solutions, only the backboning parameter *δ* is left to be determined in our pipeline. For simplicity, we focus first on a single age-sex stratum of the population (women, aged 60-69 years old) in order to demonstrate how varying *δ* impacts the scores and sub-scores of the clustering solution, and the resulting “optimal” solution i.e., the one with highest *MCS*. We chose this age stratum for illustration as it is old enough to onset multimorbidity patterns but with still a relatively low prevalence rate. Additional results from other age-sex strata can be found in the supplementary material (Figure S2).

The backboning parameter *δ* determines the degree of edge pruning that occurs in the network, with a higher value resulting in keeping only the most relevant associations. The impact of varying *δ* on each sub-score is demonstrated in Figure 2. The brown line (*E*) shows how many associations per condition on average remain after filtering, with a clear transition from a large number of edges (>50 associations/condition) when *δ<*–1, to relatively fewer edges (<10 associations/condition) when *δ>*+1. When *δ>*2, some conditions become disconnected (i.e. with no associations to any other condition within the network), and are removed from the network, reducing its size (purple line, *N*).

**Figure 2.**
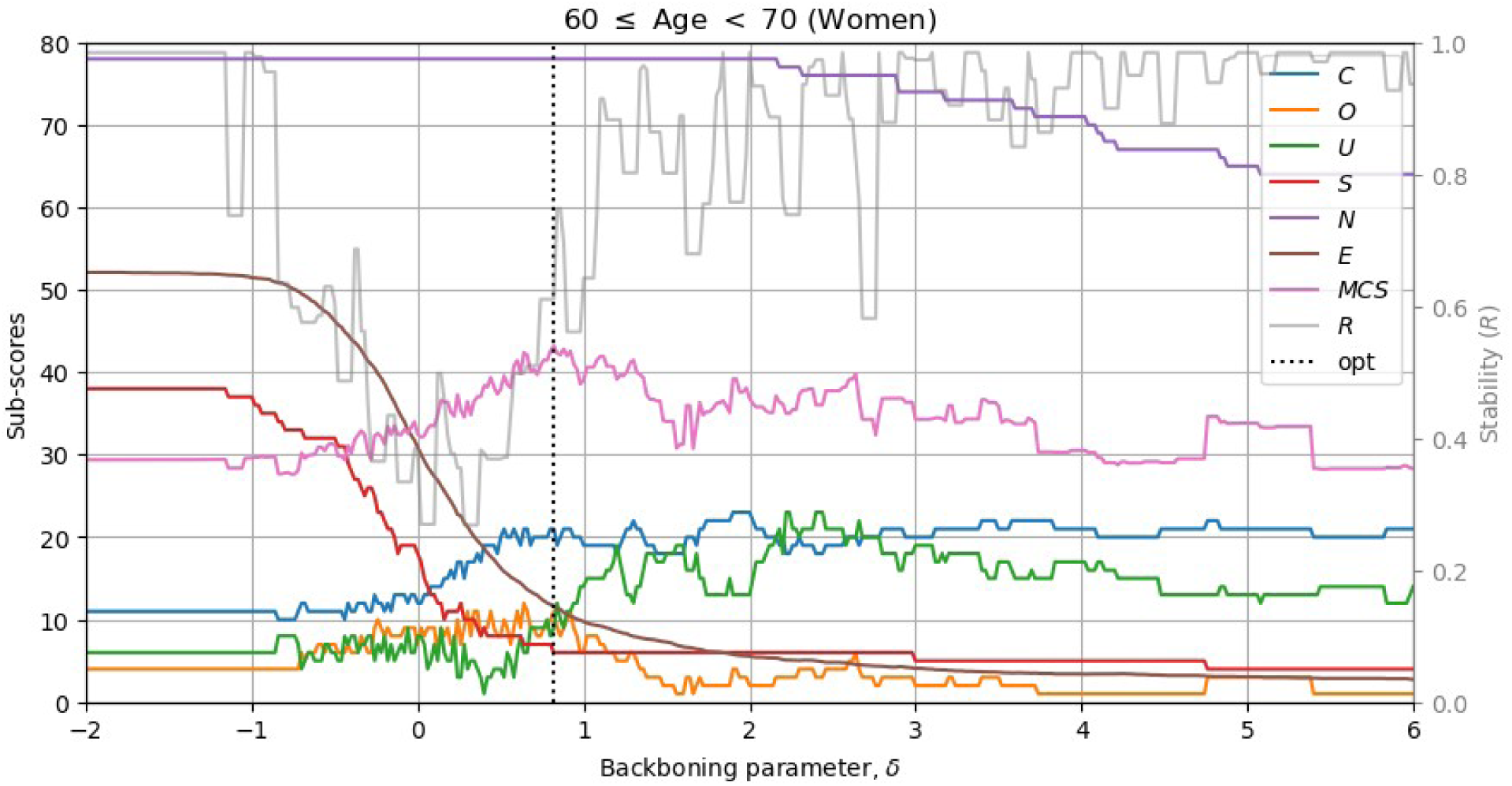
Scores of clustering solutions for 60-69 years old women for different values of the backboning parameter *δ*, in steps of 0.02, and fixed parameters *d_thresh_=1* and *cp_tresh_=0.5*. The lines represent: *C:* the number of clusters in the solution, *O:* the number of conditions with multiple memberships, *U*: the number of conditions not belonging to any cluster, *S:* the size (number of conditions) of the biggest cluster, *N:* The number of nodes in the network, *E:* the average number of connections per node, *MCS:* the *Multimorbidity Clustering* Score*, R:* the stability sub-score, *opt:* the backboning parameter *δ* that results in the highest *MCS.*

The number of clusters present in the solution increases with *δ* (blue line, *C*). At a lower *δ*, the clustering solution includes around 10 clusters, with the biggest one spanning more than half of the network (red line, *S*), and fewer than 10 conditions are not assigned to any cluster (green line,*<u>U</u>*). At higher *δ*, more than double the number of clusters are found (∼20, blue line, *C*) with the largest cluster comprising fewer than 10 conditions (red line, *S*). However, the number of conditions that are not assigned to any cluster (but still retaining an association with at least one other condition) also increases to between ∼10-20 (green line, *U*).

Varying the backboning parameter *δ* between −1 and +1 results in the greatest change in the values of all sub-scores, with more consistent values being present above and below this transition range. The stability sub-score *R* (grey line) reflects the similarity between clustering solutions for consecutive *δ*, also shows higher stability is reached when *δ* is outside of the [-1, +1] range. This means that a clustering solution with *δ* within this transition region may be problematic, as the clusters found may vary significantly with small changes in its value. Consequently, the highest *MCS* (pink line) is located at the upper end of the transition region (*δ≈*+1), where *R* is high while the trade-off between the number (*C*) and size (*S*) of clusters and the number of unclustered nodes (*U*) is optimised.

Results are relatively similar for other age-sex strata, except that the transition region widens in younger age groups (see supplementary material, Figure S2). The clustering solutions with highest *MCS* are relatively similar across age groups, as shown by their scores (Figure 3a), despite optimal values being found for different values of the backboning parameter *δ* (Figure 3b).

**Figure 3.**
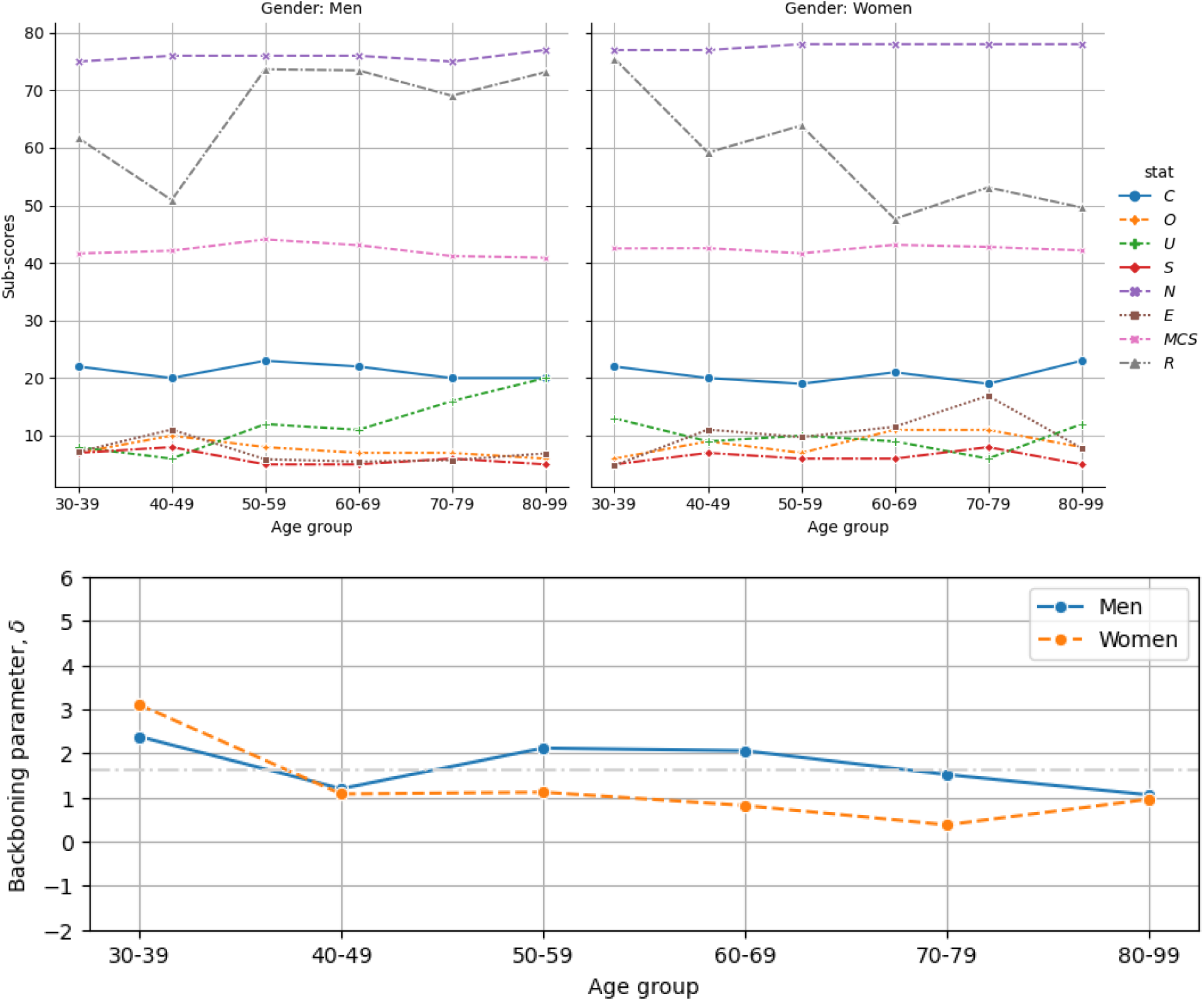
Solutions with highest *MCS* across age and sex strata. a) Sub-scores of the solutions, with lines representing scores as described in Figure 2. b) Backboning parameter *δ* values of the solutions. The backboning parameter *δ* was varied within the range [-2, 6] in steps of 0.02, while clustering parameters were fixed at *d_thresh_=1* and *cp_tresh_=0.5*.

Figure 4a provides a visualisation of the clustering solution with highest *MCS* for women aged 60-69. There are 21 clusters, comprising 69 conditions (nine conditions do not belong to any cluster and one condition has no significant associations to other conditions and is excluded from the network). Of the conditions contributing to clusters, 10 belong to two clusters, and one (skin cancer) belongs to three clusters.

**Figure 4.**
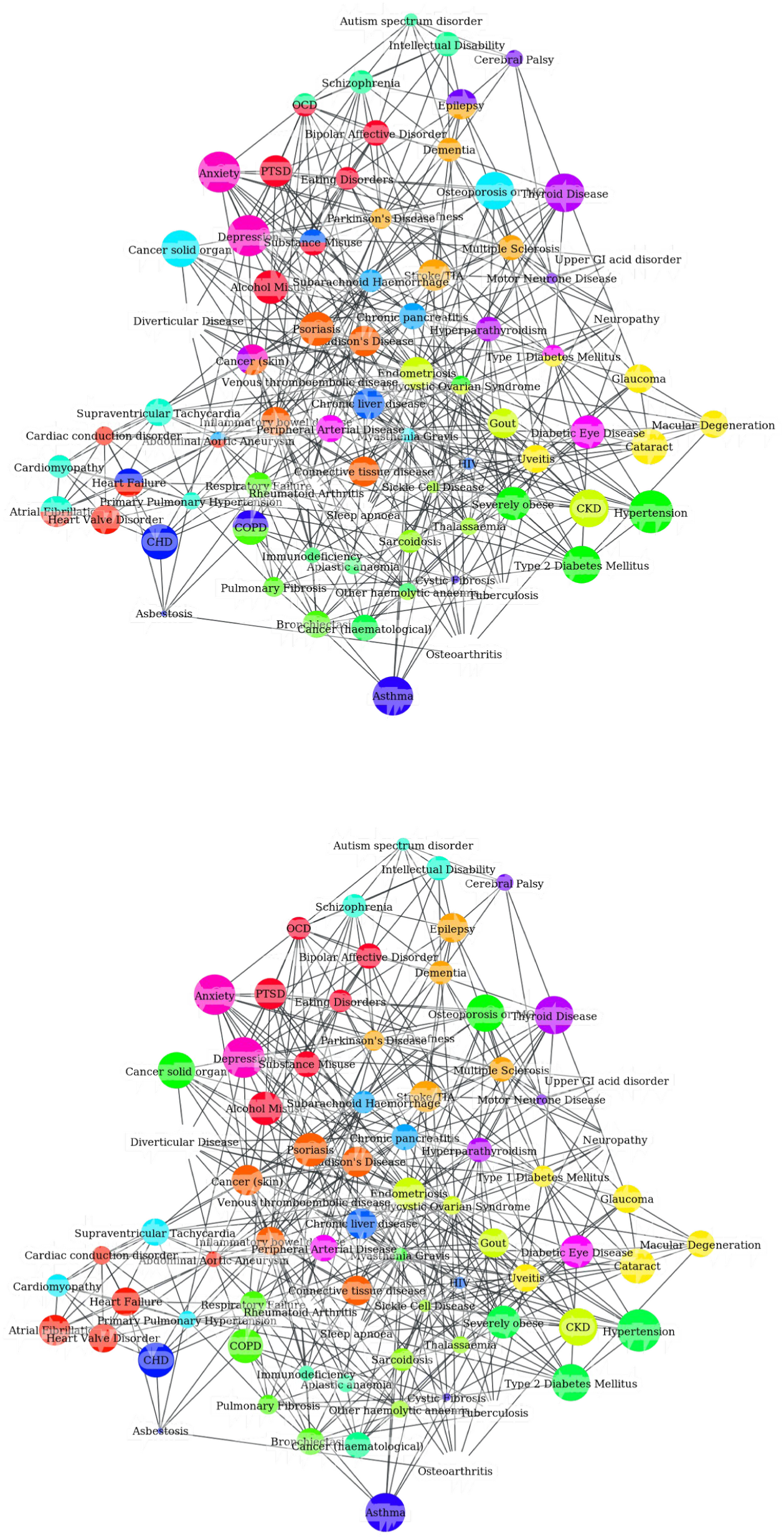
Visualisation of clustering solutions with highest *MCS* in a single age-sex stratum (women, 60-69 years old). (a) Clustering solution when allowing multiple clustering membership. The backboning parameter *δ* for this solution is 0.82. (b) Clustering solution with the same *δ* but with a version of the algorithm that only allows conditions to belong to a single cluster. Node sizes are proportional to condition prevalence.

To enhance interpretability when comparing clustering solutions across age and sex strata, Figure 5 shows specific, focused areas of the network centred on type 1 diabetes. In these focused visualisations, we show the nodes and edges for the cluster(s) that the specific condition is part of for each age and sex stratum. For example, in women aged 50-59 years old, type 1 diabetes clusters with four other conditions (diabetic eye disease, glaucoma, cataracts and macular degeneration), whereas in men aged 50-59 years old, it only clusters with two other conditions (macular degeneration and hyperparathyroidism). Similarly, across age strata, type 1 diabetes clusters with different conditions. In men in age strata 40-49 and 60-69, type 1 diabetes is a member of two distinct clusters, whereas in the other four age strata it is a member of a single cluster.

**Figure 5.**
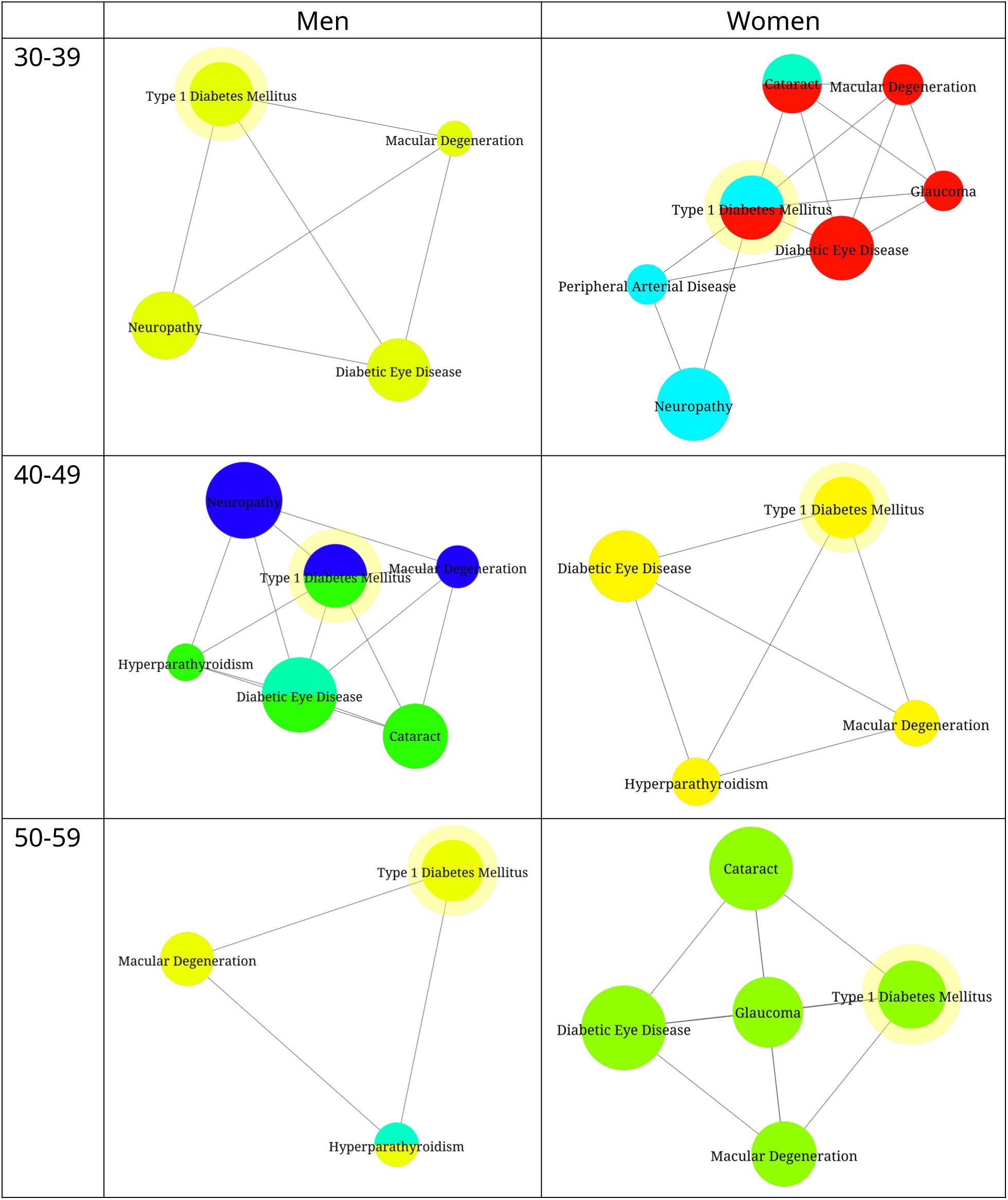
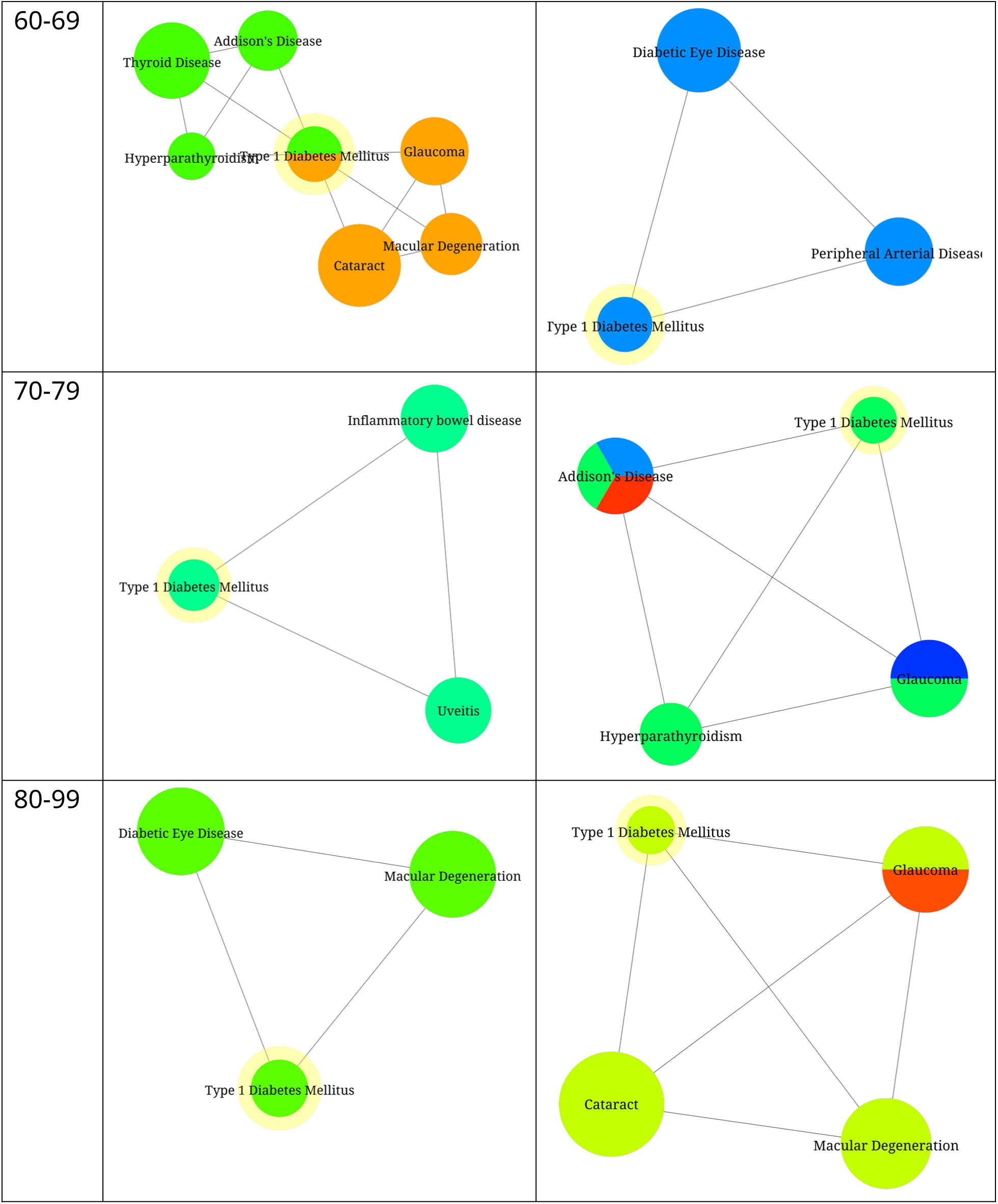
Network focus for type 1 diabetes across age-sex strata where only conditions sharing a cluster with Diabetes Type 1 (highlighted in yellow) are included. Within each network visualisation, node sizes proportional to condition prevalence, and colours are not comparable across networks.

As an alternative to condition-specific visualisations, we present a visualisation method focused on body systems (cardiovascular disease and mental health, Figure 6) to enhance interpretability. This allows comparison of how the same groups of conditions within each body system, and linked clusters, vary across age and sex strata.

**Figure 6.**
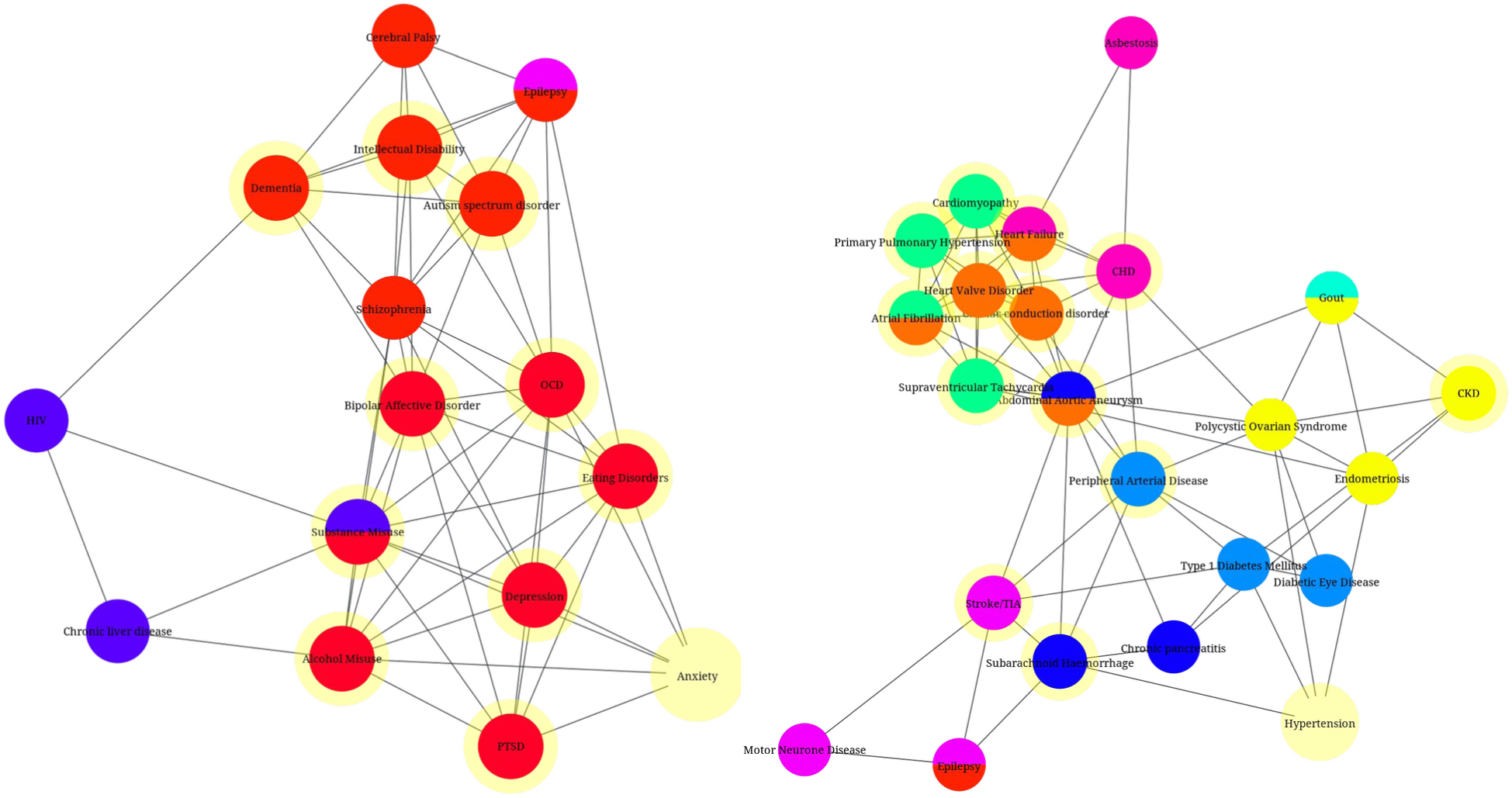
Body system focus in women 60-69 years old. a) Cardiovascular: all clusters involving any of the following conditions: abdominal aortic aneurysm, atrial fibrillation, cardiomyopathy, cardiac conduction disorder, chronic kidney disease, coronary heart disease, heart failure, heart valve disorder, hypertension, peripheral arterial disease, primary pulmonary hypertension, stroke/TIA, subarachnoid haemorrhage, supraventricular tachycardia). b) Mental health: all clusters involving any of the following conditions: alcohol misuse, anxiety, autism, bipolar affective disorder, dementia, depression, eating disorders, intellectual disability, obsessive compulsive disorder, post-traumatic stress disorder, substance misuse). Within each network visualisation, node sizes are proportional to condition prevalence.

### Comparing single with multiple membership clustering solutions

In addition to the parameters described above which determine clustering solutions, a further decision is required as to whether conditions are allowed to be members of multiple clusters. Multiple membership clustering solutions allow single conditions to belong to more the one cluster. In contrast, single membership solutions constrain conditions to belong to a single cluster. Figure 4 contrasts single and multiple membership clustering solutions in women aged 60-69 years old. Both solutions have 21 clusters, of which nine are composed of identical clustered conditions (see supplementary data S5 associated with the figure.) However, in the multiple membership solution, 12 clusters differ from the single membership solution due to having one additional condition which is present in at least two clusters. For example, there is a cluster in the single membership solution that comprises peripheral arterial disease and diabetic eye disease. In the multiple membership solution, the equivalent cluster includes an additional condition, type 1 diabetes mellitus. Similarly, chronic liver disease and HIV cluster in both solutions, but the multiple membership solution also includes substance misuse. In both examples, allowing multiple membership includes a condition which provides a potential aetiological link between the other two conditions in the cluster. A comparison of the sub-scores for solutions with other values of *δ* can be found in the supplementary material (Figure S4).

### Solutions with ‘open’ clusters

We present here results that allow for ‘open’ clusters, i.e., nodes in the clusters do not necessarily have direct associations to every other node within the cluster. This is achieved when the density threshold (*d_thresh_*) is allowed to vary within the [0, 1] range. This results in a 2D space of parameter choices, formed by the backboning parameter *δ* and the density threshold *d_thresh_*. We perform experiments across the 2D parameter space and observe the scores from the clustering solutions at different parameter combinations.

Figure 7a shows how varying *d_thresh_*while keeping *δ* fixed at three specific values (0.9, 2.1, 2.2) affects the network and clustering sub-scores. We have chosen these illustrative three *δ* values because they result in three very different *d_thresh_*values that lead to the highest *MCS* (*d_thresh_*=0.93, 0.21, and 0.64, respectively), showing the importance of accounting for the joint effect and interaction of parameters. As the *δ* is a parameter of the clustering algorithm, the sub-scores that describe the network do not change from varying this parameter, hence the number of nodes (*N*, purple line) and edges (*E*, brown line) remain constant.

**Figure 7.**
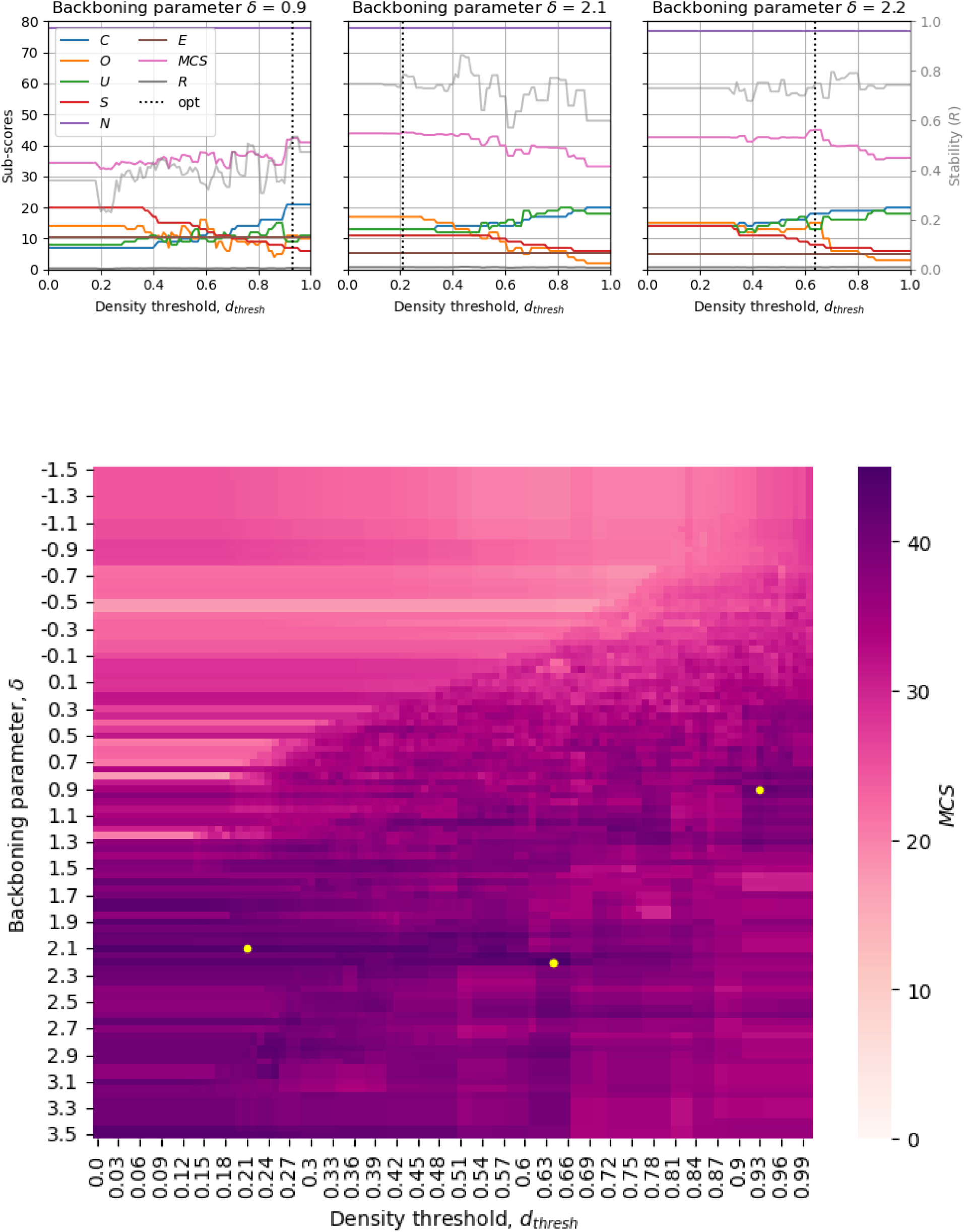
Clustering scores for different parameter combinations when clusters are allowed to be ‘open’ (i.e. with nodes not necessarily directly associated to every other node) in women aged 60-69. (a) Clustering scores for different values of *d_thresh_* and three backboning parameter *δ* values: the one leading to the highest *MCS* (*δ* =*2.2*) and two other values (*δ* =*1.9* and *δ=2.1*). Lines represent the scores as described in Figure 2. (b) Multimorbidity Clustering Score *(MCS)* values for a grid of parameter combinations *δ-d_thresh_*. Yellow dots mark the parameter combination that leads to the highest *MCS* for each of the three *δ* values in (a).

We can observe that, in all cases, lower values of the density parameter result in fewer clusters (*C*, blue line), fewer conditions unclustered (*U*, green line), a larger largest cluster (*S*, red line) and more conditions belonging to multiple clusters (*O*, orange line), although we note that the values of these sub-scores do not vary widely across the range of density thresholds, and therefore the *MCS* does not vary significantly either (pink line). Still, the *MCS* of the best parameter combinations shown in these three cases are relatively similar (42.4, 44.2, and 45.0, respectively), and therefore similarly valid solutions (according to our criteria) with different characteristics may co-exist, i.e., solutions with low backboning parameter *δ* (more edges) and *d_thresh_* close to one (closed clusters) and solutions with high *δ* (fewer edges) and low-to-moderate *d_thresh_*(open clusters).

A heatmap showing different regions of similar *MCS* values can be seen in Figure 7b. Heatmaps including the distribution of all sub-scores across the parameter space are shown in the supplementary material (Figure S6).

A visualisation of the ‘open’ clustering solution with highest *MCS* can be seen in Figure 8. This solution differs substantially to the one with highest *MCS* for ‘closed’ clusters (Figure 4a): the open solution results in 18 clusters in comparison to 21 clusters in the closed solution; the open solution results in 13 conditions in the network that do not cluster in contrast to nine in the closed solution. Only one cluster is consistent between the two solutions (Glaucoma, Type 1 Diabetes Mellitus, Uveitis, Macular Degeneration, Cataract).

**Figure 8.**
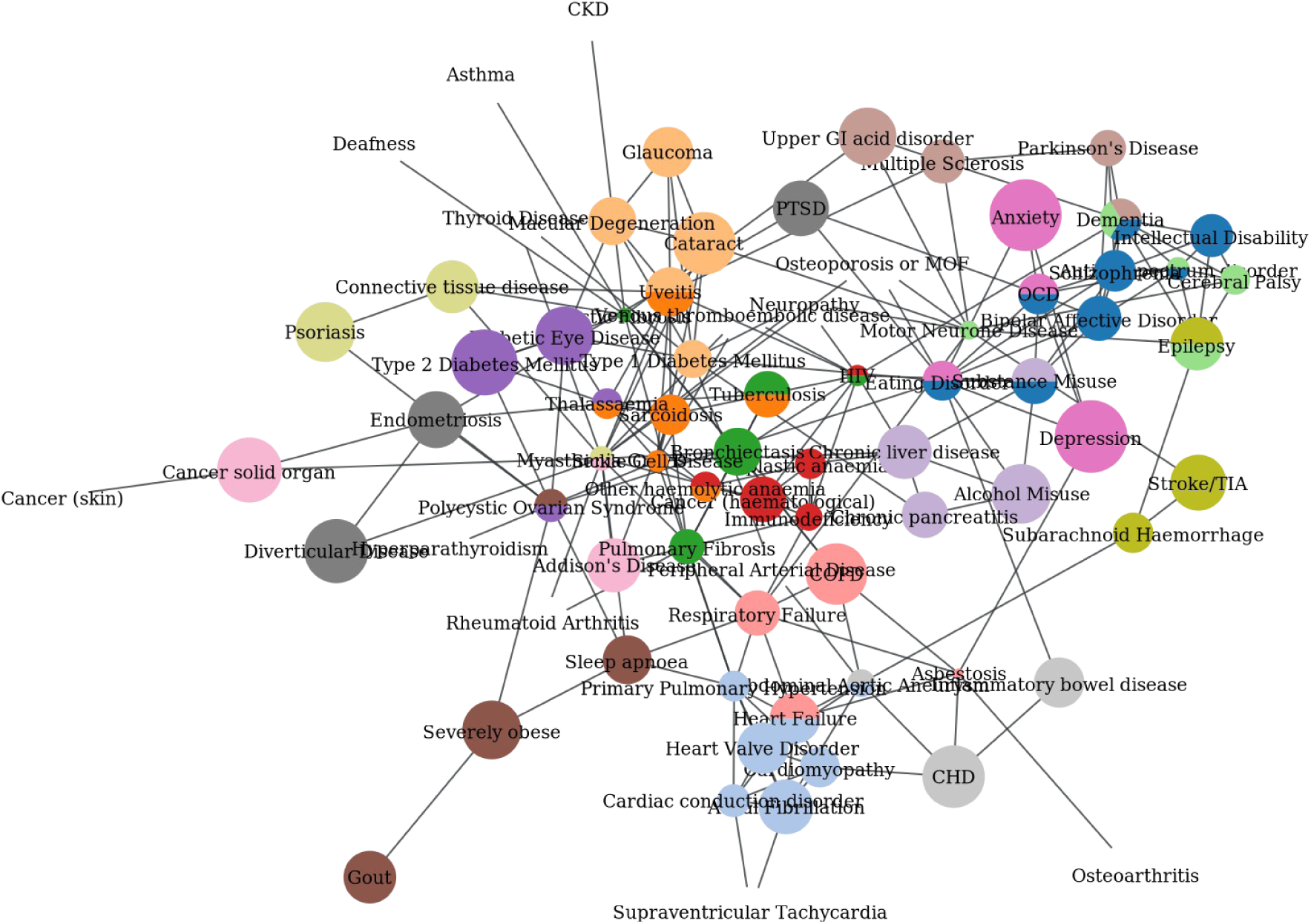
‘Open’ clustering solution with highest *MCS* in women aged 60-69, corresponding to parameter values *δ=*2.2, *d_thresh_*=0.64. A list of the conditions belonging to each cluster can be found in supplementary material S11.

As in the experiments with ‘closed’ clusters, the scores of the ‘open’ clustering solutions with highest MCS are generally similar across age groups (Figure 9), except for the appearance of a significant shift in *d_thresh_* in women 70-79 and 80-99 years old. This is a result of small variations in the *MCS* values between similarly valid solutions (supplementary material, Figure S7).

**Figure 9.**
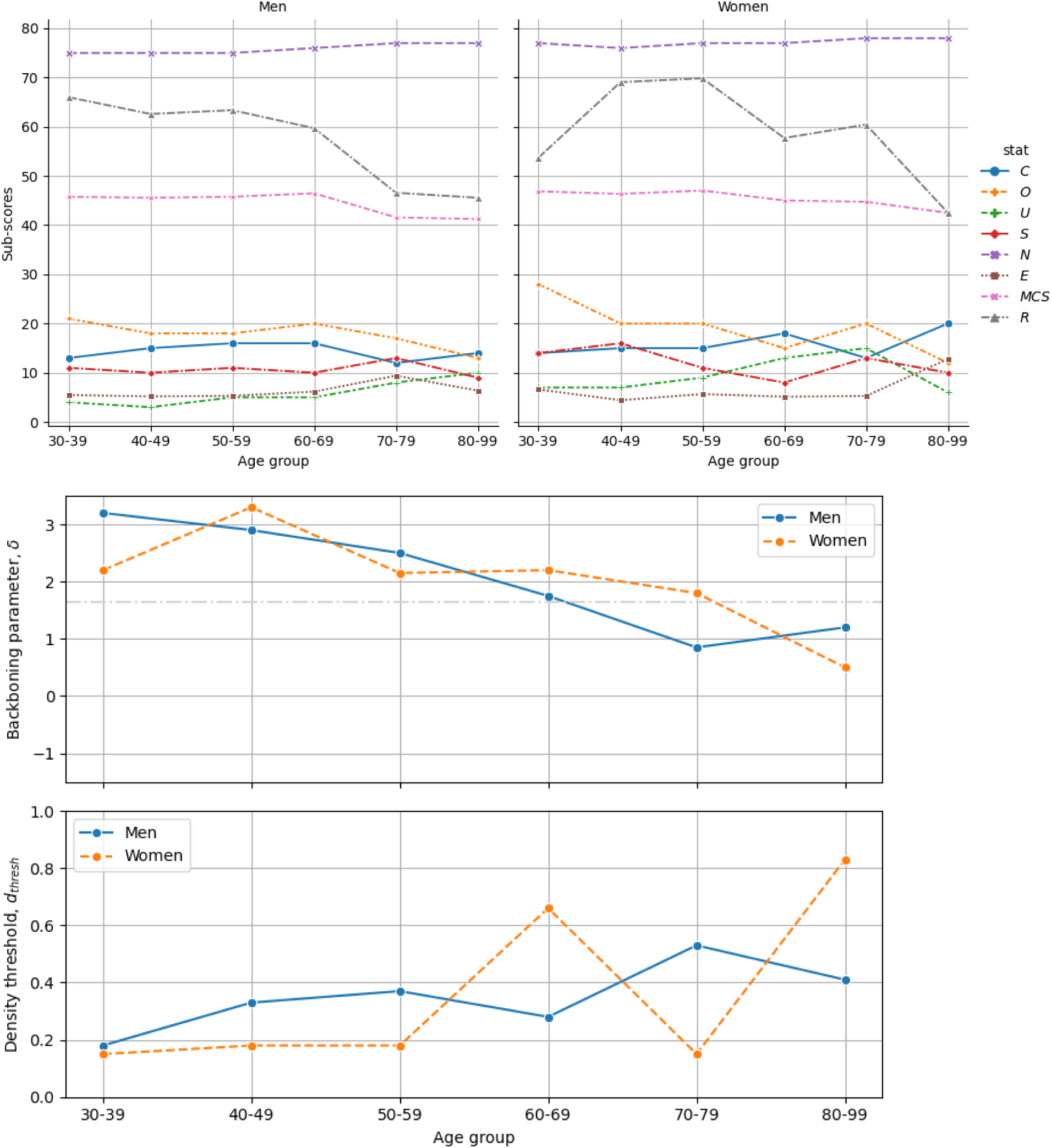
a) Sub-scores of the solutions with highest *MCS* across age and sex, and b) backboning parameter *δ* and density threshold *d_thresh_*values corresponding to the points in a). *δ* is varied within the range [-1.5, +3] in steps of 0.05 while *d_thresh_* was varied within the range [0, 1] in steps of 0.01. Lines in a) represent the scores as described in Figure 2.

## Discussion

Our study demonstrates the multiple decisions that are needed to arrive at a clustering solution using one particular clustering algorithm. At each stage, a different choice may lead to a different clustering solution. Even when applying an objective, reproducible method of selecting a solution – our proposed *Multimorbidity Clustering Score* – it remains possible to arrive at two different solutions with similar values of the *MCS.* The application of clustering methods in the field of multimorbidity research is increasing. Our study emphasises the need for transparent reporting at each stage of the development and application of a clustering solution to maximise understanding and improve adherence to reproducible research principles. This is of particular importance for any clustering solution which may translate to or be implemented in clinical contexts [34].

The intended application of a clustering algorithm should influence not only the methodological approach to clustering but also the approach to evaluate the “optimal” clustering solution [20]. However, even when the application of the clustering algorithm is known, there is currently no standardised method of determining the optimal solution [15]. We have proposed the *MCS* and sought its highest values as a way to optimise parameters, which we had prespecified in the context of our stated aim as well as preferred “statistical” properties of a solution (e.g. avoiding one large cluster as the optimal solution).

This process of prespecifying the features of a “good” clustering solution and aligning this with intended application is a key component of transparent reporting [35]. Importantly, the stability of a solution to small variations in one parameter is inherently attractive for clinical applications of clustering [19].

However, uncertainty remains as to whether these optimised parameters ultimately provide the desired clustering partitions. In particular, we demonstrate that solutions with similar *MCS* to the highest value may result in significantly different clustering solutions. This problem is further exacerbated with clustering algorithms for which independent runs may arrive at different solutions (i.e., non-deterministic) even with identical parameters, potentially pointing to ‘equally valid’ groups of solutions. While there has been recent methodology to identify and find a consensus between groups of solutions [25,36,37], further research would be required to evaluate parameter optimisation in the context of known causal frameworks and refine the score to break the ties between disparate solutions.

Application of clustering methods to identify patterns of co-occurrence of long-term conditions may reveal novel aetiological insights, indicating future mechanistic avenues of investigation [13]. In addition, condition clusters may be predictive of clinically relevant outcomes, enabling risk stratification or targeted interventions [14]. For the former application, causal inference frameworks in epidemiological research usually focus on a single association between an exposure and outcome, explicitly considering potential confounders and sources of bias, so that researchers justify the choice of study design and analytical methods to mitigate against these.

In the case of clustering algorithms, clusters are identified via computational approaches that use a number of parameters, such as the magnitude and strength of associations between individual conditions, and their “distance” from other conditions and clusters. Many hundreds or thousands of these associations are simultaneously evaluated as part of this process. However, the use of these methods does not obviate the need for grounding analyses in causal frameworks nor diminish the need for careful consideration underlying sources of bias in relation to each association, particularly when the chosen application is to surface potential aetiological mechanisms.

The ‘closed’ clustering solution, which requires all nodes within a network to be connected, removes conditions from clusters that are not directly associated with each other, and therefore should theoretically reduce the risk of conditions clustering that are less likely to have common aetiologies. For example, we found that the closed clustering solution grouped polycystic ovarian syndrome with clinically relevant conditions (hypertension, type 2 diabetes mellitus, severe obesity), whereas it clustered with thalassaemia in the open clustering solution. In addition, clustering solutions that permit conditions to have membership of multiple clusters rather than a single cluster should better reflect clinical reality [38]. Both of these choices applied to clustering algorithms are likely to reduce some non-causal associations and consequent clusters. Further consideration is required before expending additional research efforts to use clusters to discover potential novel common aetiological factors.

The literature on multimorbidity clusters uses a variety of diverse methods that result in markedly different solutions [4,5,23], while the choice of methodology and evaluation often lacks justification and alignment with the goals of the study [24].

For instance, factor analysis and multiple correspondence analysis generate ‘open’ clusters that often include negative loadings and may be more adequate for clinical management than discovery of causal mechanisms [23]. Other methods such as hierarchical clustering or latent class analysis do not assume that conditions can belong to multiple clusters and therefore may also be limited when investigating aetiology, where multiple causal pathways are often intertwined.

Importantly, each of these algorithms implicitly defines what a cluster is and uses some metric to qualify, rate, and choose among solutions resulting from variations in its parameters [20]. Researchers applying these methods to the multimorbidity context should be aware of these assumptions and explicitly align them with their research goals [35].

Our study has a number of strengths. We have provided a detailed analysis pipeline to demonstrate the impact of decisions at each step in implementing a clustering algorithm to multiple long-term conditions. We have also provided one method of identifying optimal clustering solutions using a range of objective scores. However, we do not propose that the score would be universally valid for all methods applied to long-term condition clustering, as its specification also takes into account the methodological pipeline used. On the contrary, we propose it as a guideline that can further be adapted to the particularities of other approaches. In addition, we recognise the difficulties in interpreting whole network visualisation of clusters, and have therefore provided single condition and body system focused views of areas of the network to aid clinical understanding.

In addition, we have used real world data relating to over 7 million individuals registered with GP practices in England, reducing selection bias and improving generalisability. Our pipeline is tailored to the discovery of causal mechanisms, for which we have chosen methods that correct for co-occurrence by chance, allow for multiple cluster membership, and can generate ‘closed’ clusters.

However, our study has limitations. Whilst real-world routine healthcare data has advantages over simulated data, diagnostic coding in electronic health records are prone to error and bias [39]. The impact of this can be partly mitigated through incorporating expert knowledge of the processes involved in data collection and data entry in primary care (our investigator group included primary care practitioners) in the derivation of code lists for long-term conditions from data sources. In addition, the cross-sectional nature of the dataset limited our ability to understand longitudinal long-term condition clusters [6,10]. We used stratified analyses which reduced the confounding effect of age and sex on associations between conditions [40]. However, this does not account for other potential confounders, such as social deprivation or ethnicity, which my contribute to clustering of conditions [41].

In addition, the ABC method of identifying associations has intrinsic ceiling effects that limit the appearance of associations between some common conditions (e.g. chronic heart disease, hypertension), particularly in older age groups [22]. This is because of the weaker signals and limited capability of inducing these ‘from the data alone’. However, the aim of our study was to explore and discover possible new groups of associations from routine data, which is particularly advantageous as a method for findings between rarer conditions or combinations.

Decisions such as whether to allow open vs closed clusters, or whether individual conditions can belong to more than one cluster, result in different clustering solutions. Such decisions are primarily informed by domain-expertise (drawing from epidemiological principles and clinical medicine respectively), rather than related to optimising algorithm parameters. Similarly, deciding which features of clustering solutions are desirable is largely driven by the intended application of the solution.

For these reasons, researchers planning to use clustering methods in healthcare contexts should provide a clear justification for each step in the decision-making process. Furthermore, transparency and reproducibility of healthcare research have been enhanced by the development and increasing use of reporting research guidelines for a variety of study designs and applications, including randomised controlled trials [42], epidemiological studies [43] and prediction models [44]. Our study highlights potential components of an equivalent reporting guidelines for studies using clustering methods.

Using electronic health records for 7 million patients in England, we have demonstrated the importance of transparently reporting the decisions required to develop clustering solutions for research focussing on long-term conditions. We have provided a framework to specify and quantify the desirable features of an optimal clustering solution, allowing studies using clustering methods to be reproduced by other researchers. Despite the advancement and increasing addition of data-driven approaches, further work is needed to develop rigorous methodology tailored for long-term condition clustering and to improve transparent reporting of studies. This will bring us closer to realising the benefits for people living with multiple long-term conditions.

## Data availability

The data that support the findings of this study are available from the Clinical Practice Research Datalink (CPRD) but restrictions apply to the availability of these data, which were used under licence for the current study, and so are not publicly available. Data are however available from the authors upon reasonable request and with permission of the Clinical Practice Research Datalink (CPRD) (https://www.cprd.com/).

## Acknowledgements

The study was funded by the National Institute for Health and Care Research (NIHR) Artificial Intelligence and Multimorbidity: Clustering in Individuals, Space and Clinical Context (NIHR202639). The funder had no role in conduct of the study, interpretation, or the decision to submit for publication. The views expressed are those of the authors and not necessarily those of the NIHR, the Department of Health and Social Care.

This study is based in part on data from the Clinical Practice Research Datalink obtained under licence from the UK Medicines and Healthcare products Regulatory Agency. The data is provided by patients and collected by the NHS as part of their care and support. The interpretation and conclusions contained in this study are those of the author/s alone.

