## Supplementary Materials for "Towards reproducible multimorbidity clustering in electronic health records: a transparent pipeline for aligning research aims and methodology"

* Equal contribution

#### S1. Study population demography and prevalence of long-term conditions

| | **Men**<br>**No. (%) of men**<br>**(N=3733207)** | **Women**<br>**No. (%) of women**<br>**(N=3757667)** |
|---|---|---|
| Age-group | | |
| 30-39 | 841765 (22.55%) | 806156 (21.45%) |
| 40-49 | 809411 (21.68%) | 758652 (20.19%) |
| 50-59 | 813714 (21.80%) | 773995 (20.60%) |
| 60-69 | 588217 (15.76%) | 588123 (15.65%) |
| 70-79 | 438586 (11.75%) | 482340 (12.84%) |
| 80-99 | 241514 (6.47%) | 348401 (9.27%) |
| Hypertension | 937570 (25.11%) | 943159 (25.10%) |
| Depression | 453233 (12.14%) | 808372 (21.51%) |
| Anxiety | 453755 (12.15%) | 801713 (21.34%) |
| Osteoarthritis | 493687 (13.22%) | 722018 (19.21%) |
| Asthma | 498276 (13.35%) | 616713 (16.41%) |
| Chronic kidney disease | 334523 (8.96%) | 436280 (11.61%) |
| Deafness | 326845 (8.76%) | 312023 (8.30%) |
| Type 2 Diabetes Mellitus | 334481 (8.96%) | 269652 (7.18%) |
| Cataract | 237256 (6.36%) | 336635 (8.96%) |
| Coronary heart disease | 328166 (8.79%) | 223384 (5.94%) |
| Osteoporosis or MOF | 171450 (4.59%) | 366384 (9.75%) |
| Thyroid Disease | 95505 (2.56%) | 390720 (10.40%) |
| Diverticular Disease | 196363 (5.26%) | 243995 (6.49%) |
| Cancer solid organ | 185125 (4.96%) | 240370 (6.40%) |
| Alcohol Misuse | 263808 (7.07%) | 152875 (4.07%) |
| Psoriasis | 156620 (4.20%) | 161260 (4.29%) |
| Atrial Fibrillation | 171725 (4.60%) | 131367 (3.50%) |
| Chronic Obstructive Pulmonary Disease | 150910 (4.04%) | 147419 (3.92%) |
| Gout | 233934 (6.27%) | 62013 (1.65%) |
| Benign Prostatic Hyperplasia | 274172 (7.34%) | 120 (0.00%) |
| Diabetic Eye Disease | 143512 (3.84%) | 109917 (2.93%) |
| Upper GI acid disorder | 137962 (3.70%) | 103434 (2.75%) |
| Stroke/TIA | 121658 (3.26%) | 112103 (2.98%) |
| Neuropathy | 111606 (2.99%) | 112178 (2.99%) |
| Severely obese | 62648 (1.68%) | 135418 (3.60%) |
| Venous thromboembolic disease | 80428 (2.15%) | 102804 (2.74%) |
| Cancer (skin excl. melanoma) | 95311 (2.55%) | 85499 (2.28%) |

| | | |
|---|---|---|
| Heart Failure | 97853 (2.62%) | 77003 (2.05%) |
| Post-traumatic stress disorder | 63277 (1.69%) | 94549 (2.52%) |
| Heart Valve Disorder | 77411 (2.07%) | 76465 (2.03%) |
| Substance Misuse | 100941 (2.70%) | 50312 (1.34%) |
| Epilepsy | 69861 (1.87%) | 72237 (1.92%) |
| Chronic liver disease | 75666 (2.03%) | 62063 (1.65%) |
| Endometriosis | 85 (0.00%) | 130971 (3.49%) |
| Connective tissue disease (excl. RA) | 50262 (1.35%) | 73940 (1.97%) |
| Macular Degeneration | 44058 (1.18%) | 70142 (1.87%) |
| Glaucoma | 52998 (1.42%) | 59230 (1.58%) |
| Rheumatoid Arthritis | 33479 (0.90%) | 76888 (2.05%) |
| Dementia | 40678 (1.09%) | 64882 (1.73%) |
| Peripheral Arterial Disease | 61771 (1.65%) | 38157 (1.02%) |
| Sleep apnoea | 66217 (1.77%) | 26990 (0.72%) |
| Inflammatory bowel disease | 42980 (1.15%) | 47136 (1.25%) |
| Addison's Disease | 21564 (0.58%) | 63718 (1.70%) |
| Uveitis | 36475 (0.98%) | 38011 (1.01%) |
| Tuberculosis | 30495 (0.82%) | 33118 (0.88%) |
| Supraventricular Tachycardia | 25269 (0.68%) | 36433 (0.97%) |
| Polycystic Ovarian Syndrome | 23 (0.00%) | 58348 (1.55%) |
| Cancer (haematological) | 31986 (0.86%) | 24806 (0.66%) |
| Chronic pancreatitis | 27059 (0.72%) | 28920 (0.77%) |
| Bronchiectasis | 23729 (0.64%) | 31330 (0.83%) |
| Intellectual Disability or Down Syndrome | 28922 (0.77%) | 22602 (0.60%) |
| Respiratory Failure | 22374 (0.60%) | 23266 (0.62%) |
| Bipolar Affective Disorder | 17309 (0.46%) | 26451 (0.70%) |
| Obsessive Compulsive Disorder | 14917 (0.40%) | 21212 (0.56%) |
| Schizophrenia | 20128 (0.54%) | 15981 (0.43%) |
| Cardiomyopathy | 20903 (0.56%) | 11237 (0.30%) |
| Type 1 Diabetes Mellitus | 17673 (0.47%) | 12897 (0.34%) |
| Abdominal Aortic Aneurysm | 22481 (0.60%) | 5635 (0.15%) |
| Eating Disorders | 1489 (0.04%) | 26341 (0.70%) |
| Parkinson's Disease | 15496 (0.42%) | 11297 (0.30%) |
| Cardiac conduction disorder | 15778 (0.42%) | 10688 (0.28%) |
| Hyperparathyroidism | 6609 (0.18%) | 18268 (0.49%) |
| Multiple Sclerosis | 6901 (0.18%) | 17366 (0.46%) |
| Sarcoidosis | 10601 (0.28%) | 10533 (0.28%) |
| Subarachnoid Haemorrhage | 8324 (0.22%) | 10185 (0.27%) |
| Pulmonary Fibrosis | 9502 (0.25%) | 7764 (0.21%) |
| Autism spectrum disorder | 8899 (0.24%) | 3304 (0.09%) |

| | | |
|---|---|---|
| Other haemolytic anaemia | 4085 (0.11%) | 5494 (0.15%) |
| Primary Pulmonary Hypertension | 3892 (0.10%) | 5641 (0.15%) |
| Cerebral Palsy | 5042 (0.14%) | 4339 (0.12%) |
| Thalassaemia | 2973 (0.08%) | 6142 (0.16%) |
| Asbestosis | 7554 (0.20%) | 460 (0.01%) |
| Aplastic anaemia | 4078 (0.11%) | 3878 (0.10%) |
| HIV | 4322 (0.12%) | 2446 (0.07%) |
| Immunodeficiency | 2050 (0.05%) | 2527 (0.07%) |
| Sickle Cell Disease | 1451 (0.04%) | 2531 (0.07%) |
| Myasthenia Gravis | 1846 (0.05%) | 1861 (0.05%) |
| Motor Neurone Disease | 1138 (0.03%) | 787 (0.02%) |
| Cystic Fibrosis | 452 (0.01%) | 598 (0.02%) |

Conditions listed in descending order of prevalence in combined population of men and women. MOF – major osteoporotic fracture; TIA – transient ischaemic attack; RA – rheumatoid arthritis; HIV – Human Immunodeficiency Virus. Conditions with zero cases for men or for women are specific to the other sex.

| | All participants (both women and men) (N=7490874) | 30-39 % with LTC N= 1647921 | 40-49 % with LTC N= 1568063 | 50-59 % with LTC N= 1587709 | 60-69 % with LTC N= 1176340 | 70-79 % with LTC N= 920926 | 80-99 % with LTC N= 589915 |
|---|---|---|---|---|---|---|---|
| Hypertension | 1880729 (25.11%) | 1.98 | 7.8 | 20 | 37.91 | 56.92 | 74.29 |
| Depression | 1261605 (16.84%) | 12.91 | 17.03 | 19.35 | 19.02 | 16.93 | 16.1 |
| Anxiety | 1255468 (16.76%) | 15.05 | 16.99 | 17.95 | 17.67 | 16.9 | 15.68 |
| Osteoarthritis | 1215705 (16.23%) | 1 | 3.89 | 11.63 | 25.14 | 38.56 | 51.31 |
| Asthma | 1114989 (14.88%) | 16.78 | 14.92 | 14.15 | 13.74 | 14.23 | 14.77 |
| Chronic kidney disease | 770803 (10.29%) | 1.66 | 2.54 | 5.05 | 10.95 | 23.39 | 47.31 |
| Deafness | 638868 (8.53%) | 3.35 | 3.86 | 5.62 | 9.53 | 16.25 | 29.16 |
| Type 2 Diabetes Mellitus | 604133 (8.06%) | 0.82 | 3.09 | 7.07 | 12.78 | 17.32 | 20.35 |
| Cataract | 573891 (7.66%) | 0.29 | 0.63 | 1.9 | 6.57 | 18.66 | 47.46 |

| | | | | | | | |
|---|---|---|---|---|---|---|---|
| Coronary heart disease | 551550 (7.36%) | 0.57 | 1.38 | 4.15 | 9.92 | 18.05 | 29.1 |
| Osteoporosis or MOF | 537834 (7.18%) | 3.28 | 3.1 | 4.32 | 7.86 | 13.28 | 25.73 |
| Thyroid Disease | 486225 (6.49%) | 2.42 | 4.03 | 5.89 | 8.48 | 11.12 | 14.82 |
| Diverticular Disease | 440358 (5.88%) | 0.23 | 0.92 | 3.76 | 7.82 | 14.89 | 22.58 |
| Cancer solid organ | 425495 (5.68%) | 0.66 | 1.57 | 3.74 | 7.86 | 14.16 | 18.28 |
| Alcohol Misuse | 416683 (5.56%) | 4.56 | 5.53 | 6.54 | 6.77 | 5.61 | 3.31 |
| Psoriasis | 317880 (4.24%) | 3.05 | 3.79 | 4.43 | 5.07 | 5.4 | 4.85 |
| Atrial Fibrillation | 303092 (4.05%) | 0.19 | 0.48 | 1.32 | 3.86 | 10.02 | 22.67 |
| Chronic Obstructive Pulmonary Disease | 298329 (3.98%) | 0.21 | 0.75 | 2.47 | 6.14 | 10.46 | 12.78 |
| Gout | 295947 (3.95%) | 0.62 | 1.72 | 3.38 | 5.75 | 8.17 | 10.56 |
| Benign Prostatic Hyperplasia | 274292 (3.66%) | 0.04 | 0.23 | 1.31 | 4.84 | 10.85 | 15.67 |
| Diabetic Eye Disease | 253429 (3.38%) | 0.45 | 1.2 | 2.71 | 5.17 | 7.41 | 9.32 |
| Upper GI acid disorder | 241396 (3.22%) | 0.59 | 1.33 | 2.65 | 4.61 | 6.67 | 9.03 |
| Stroke/TIA | 233761 (3.12%) | 0.27 | 0.63 | 1.56 | 3.56 | 7.28 | 14.56 |
| Neuropathy | 223784 (2.99%) | 0.8 | 1.69 | 3.01 | 4.38 | 5.36 | 5.98 |
| Severely obese | 198066 (2.64%) | 2.07 | 2.78 | 3.37 | 3.26 | 2.37 | 1.11 |
| Venous thromboembolic disease | 183232 (2.45%) | 0.73 | 1.3 | 1.91 | 2.94 | 4.66 | 7.29 |
| Cancer (skin excl. melanoma) | 180810 (2.41%) | 0.11 | 0.4 | 1.06 | 2.55 | 5.94 | 12.07 |
| Heart Failure | 174856 (2.33%) | 0.12 | 0.3 | 0.87 | 2.32 | 5.29 | 13.29 |
| Post-traumatic stress disorder | 157826 (2.11%) | 1.64 | 2.17 | 2.47 | 2.33 | 1.98 | 2.02 |
| Heart Valve Disorder | 153876 (2.05%) | 0.23 | 0.4 | 0.84 | 1.99 | 4.69 | 10.86 |

| | | | | | | | |
|---|---|---|---|---|---|---|---|
| Substance Misuse | 151253 (2.02%) | 2.95 | 2.99 | 1.95 | 1.07 | 0.73 | 0.91 |
| Epilepsy | 142098 (1.90%) | 1.56 | 1.78 | 1.99 | 2.06 | 2.11 | 2.21 |
| Chronic liver disease | 137729 (1.84%) | 0.87 | 1.65 | 2.31 | 2.63 | 2.25 | 1.56 |
| Endometriosis | 131056 (1.75%) | 1.28 | 2.42 | 2.61 | 1.67 | 0.88 | 0.51 |
| Connective tissue disease (excl. RA) | 124202 (1.66%) | 0.4 | 0.65 | 1.03 | 1.8 | 3.46 | 6.45 |
| Macular Degeneration | 114200 (1.52%) | 0.05 | 0.12 | 0.32 | 1 | 3.21 | 11.03 |
| Glaucoma | 112228 (1.50%) | 0.06 | 0.18 | 0.57 | 1.53 | 3.66 | 8.05 |
| Rheumatoid Arthritis | 110367 (1.47%) | 0.29 | 0.62 | 1.21 | 2.14 | 3.13 | 3.84 |
| Dementia | 105560 (1.41%) | 0.02 | 0.05 | 0.14 | 0.55 | 2.53 | 12.26 |
| Peripheral Arterial Disease | 99928 (1.33%) | 0.06 | 0.17 | 0.6 | 1.75 | 3.52 | 5.74 |
| Sleep apnoea | 93207 (1.24%) | 0.4 | 0.93 | 1.62 | 2.1 | 1.79 | 0.89 |
| Inflammatory bowel disease | 90116 (1.20%) | 0.8 | 1 | 1.21 | 1.46 | 1.7 | 1.58 |
| Addison's Disease | 85282 (1.14%) | 0.31 | 0.54 | 0.9 | 1.55 | 2.24 | 3.13 |
| Uveitis | 74486 (0.99%) | 0.43 | 0.72 | 1.02 | 1.3 | 1.53 | 1.81 |
| Tuberculosis | 63613 (0.85%) | 0.49 | 0.63 | 0.63 | 0.91 | 1.32 | 2.15 |
| Supraventricular Tachycardia | 61702 (0.82%) | 0.31 | 0.43 | 0.65 | 1.02 | 1.55 | 2.24 |
| Polycystic Ovarian Syndrome | 58371 (0.78%) | 1.98 | 1.17 | 0.38 | 0.08 | 0.03 | 0.01 |
| Cancer (haematological) | 56792 (0.76%) | 0.19 | 0.31 | 0.54 | 1.01 | 1.67 | 2.21 |
| Chronic pancreatitis | 55979 (0.75%) | 0.31 | 0.5 | 0.7 | 0.92 | 1.22 | 1.64 |
| Bronchiectasis | 55059 (0.74%) | 0.09 | 0.18 | 0.37 | 0.96 | 2 | 2.57 |
| Intellectual Disability or Down Syndrome | 51524 (0.69%) | 0.85 | 0.71 | 0.79 | 0.66 | 0.46 | 0.3 |

| | | | | | | | |
|---|---|---|---|---|---|---|---|
| Respiratory Failure | 45640 (0.61%) | 0.12 | 0.21 | 0.4 | 0.8 | 1.35 | 2.06 |
| Bipolar Affective Disorder | 43760 (0.58%) | 0.48 | 0.62 | 0.68 | 0.64 | 0.57 | 0.43 |
| Obsessive Compulsive Disorder | 36129 (0.48%) | 0.66 | 0.59 | 0.48 | 0.39 | 0.29 | 0.19 |
| Schizophrenia | 36109 (0.48%) | 0.41 | 0.52 | 0.56 | 0.49 | 0.43 | 0.42 |
| Cardiomyopathy | 32140 (0.43%) | 0.09 | 0.17 | 0.36 | 0.63 | 0.94 | 1.04 |
| Type 1 Diabetes Mellitus | 30570 (0.41%) | 0.44 | 0.48 | 0.48 | 0.39 | 0.27 | 0.18 |
| Abdominal Aortic Aneurysm | 28116 (0.38%) | 0.01 | 0.02 | 0.05 | 0.41 | 1.05 | 2.11 |
| Eating Disorders | 27830 (0.37%) | 0.56 | 0.56 | 0.38 | 0.21 | 0.09 | 0.04 |
| Parkinson's Disease | 26793 (0.36%) | 0.01 | 0.02 | 0.09 | 0.35 | 1.04 | 1.9 |
| Cardiac conduction disorder | 26466 (0.35%) | 0.03 | 0.04 | 0.09 | 0.24 | 0.75 | 2.45 |
| Hyperparathyroidism | 24877 (0.33%) | 0.05 | 0.11 | 0.23 | 0.43 | 0.72 | 1.21 |
| Multiple Sclerosis | 24267 (0.32%) | 0.15 | 0.3 | 0.43 | 0.47 | 0.4 | 0.21 |
| Sarcoidosis | 21134 (0.28%) | 0.09 | 0.2 | 0.35 | 0.43 | 0.43 | 0.35 |
| Subarachnoid Haemorrhage | 18509 (0.25%) | 0.06 | 0.13 | 0.25 | 0.38 | 0.46 | 0.47 |
| Pulmonary Fibrosis | 17266 (0.23%) | 0.02 | 0.03 | 0.09 | 0.24 | 0.6 | 1.13 |
| Autism spectrum disorder | 12203 (0.16%) | 0.33 | 0.18 | 0.15 | 0.09 | 0.04 | 0.02 |
| Other haemolytic anaemia | 9579 (0.13%) | 0.12 | 0.13 | 0.12 | 0.12 | 0.14 | 0.17 |
| Primary Pulmonary Hypertension | 9533 (0.13%) | 0.02 | 0.03 | 0.06 | 0.12 | 0.27 | 0.66 |
| Cerebral Palsy | 9381 (0.13%) | 0.17 | 0.14 | 0.13 | 0.12 | 0.08 | 0.05 |
| Thalassaemia | 9115 (0.12%) | 0.14 | 0.14 | 0.12 | 0.11 | 0.1 | 0.1 |
| Asbestosis | 8014 (0.11%) | 0 | 0 | 0.01 | 0.09 | 0.39 | 0.55 |
| Aplastic anaemia | 7956 (0.11%) | 0.04 | 0.06 | 0.08 | 0.14 | 0.18 | 0.28 |
| HIV | 6768 (0.09%) | 0.05 | 0.15 | 0.16 | 0.08 | 0.03 | 0.01 |
| Immunodeficiency | 4577 (0.06%) | 0.04 | 0.04 | 0.05 | 0.08 | 0.11 | 0.1 |
| Sickle Cell Disease | 3982 (0.05%) | 0.07 | 0.07 | 0.06 | 0.03 | 0.02 | 0.03 |

| | | | | | | | |
|---|---|---|---|---|---|---|---|
| Myasthenia Gravis | 3707 (0.05%) | 0.01 | 0.02 | 0.04 | 0.06 | 0.11 | 0.14 |
| Motor Neurone Disease | 1925 (0.03%) | 0.01 | 0.01 | 0.02 | 0.04 | 0.06 | 0.06 |
| Cystic Fibrosis | 1050 (0.01%) | 0.02 | 0.01 | 0.01 | 0.01 | 0.01 | 0.01 |

#### S2. Clustering sub-scores for all age-sex strata

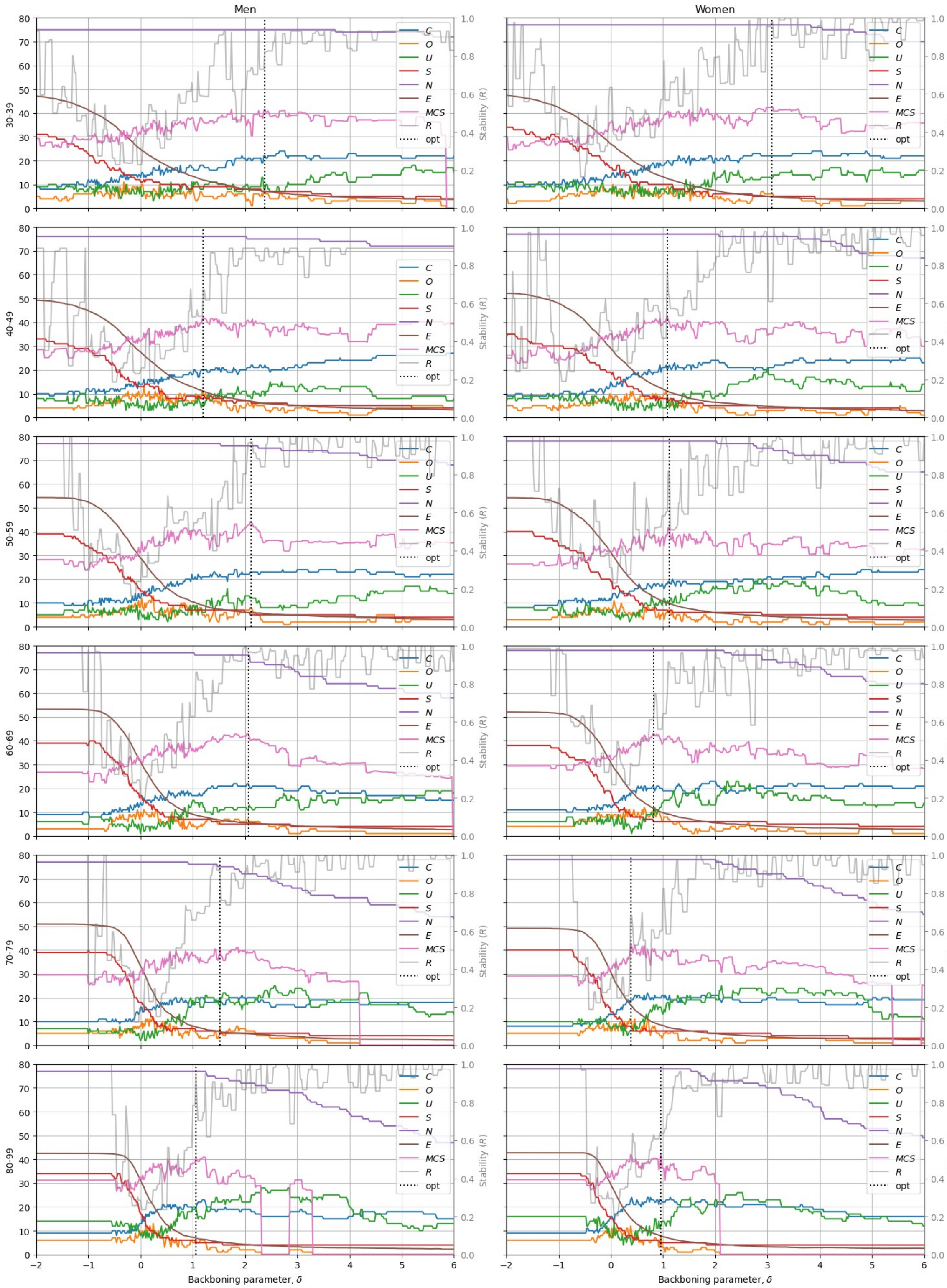


**Figure S2.** Scores of clustering solutions for all age-sex strata for different values of the backboning parameter $\delta$, in steps of 0.02, and fixed parameters $d_{thresh}$*=1* and

$cp_{tresh}$=*0.5*. The lines represent: *C:* the number of clusters in the solution, *O:* the number of conditions with multiple memberships, *U*: the number of conditions not belonging to any cluster, *S:* the size (number of conditions) of the biggest cluster, *N:* The number of nodes in the network, *E:* the average number of connections per node, *MCS:* the *Multimorbidity Clustering* Score, *R:* the stability sub-score, *opt:* the backboning parameter *δ* that results in the highest *MCS.*

#### S3. Different definitions of the Stability sub-score

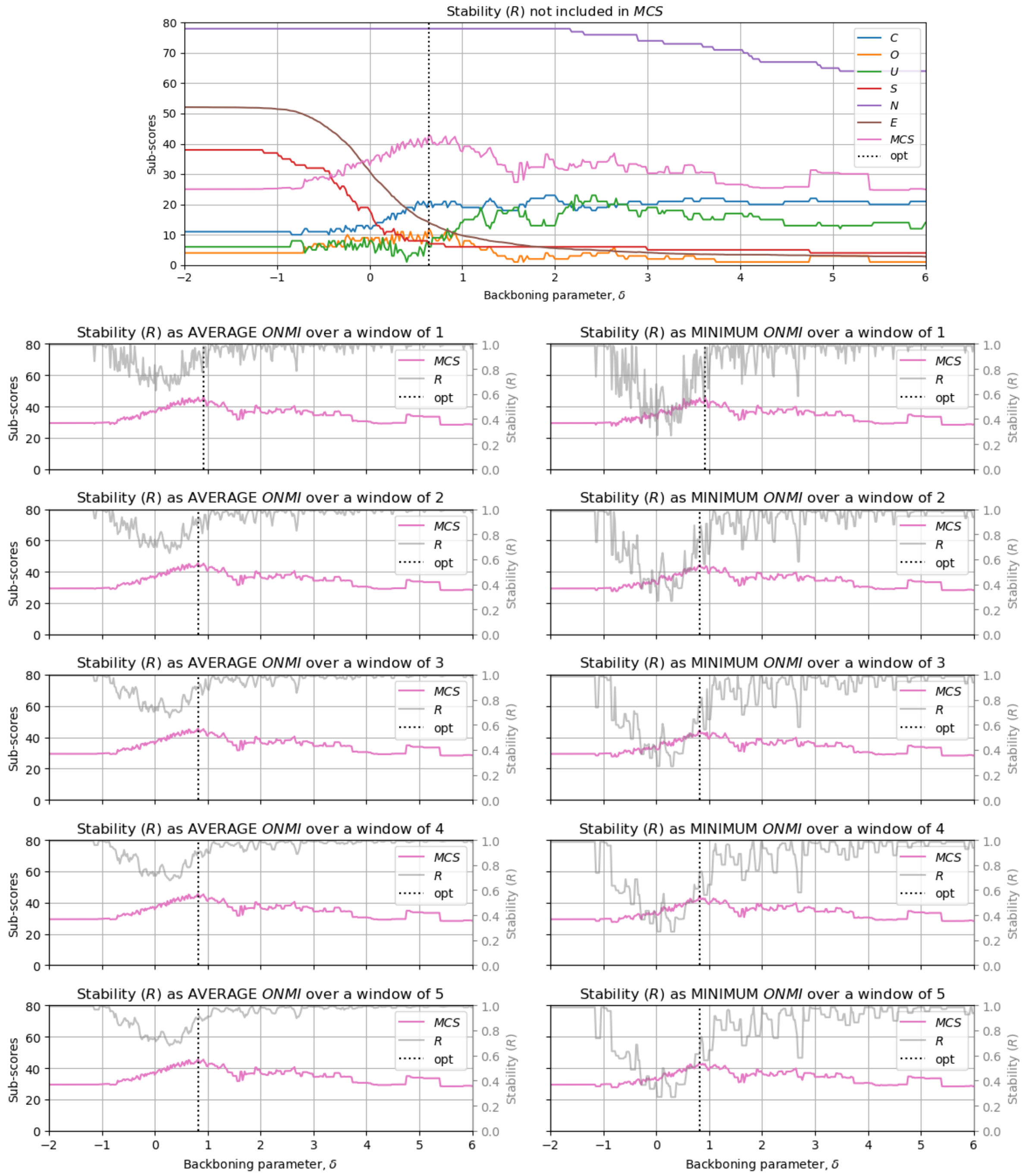


**Figure S3.** Different definitions of the stability sub-score (*R*) and the resulting *Multimorbidity Clustering Score (MCS)* of clustering solutions for 60-69 years old women for different values of the backboning parameter $\delta$, in steps of 0.02, and fixed parameters $d_{thresh}=1$ and $cp_{tresh}=0.5$.

#### S4. Single and multiple cluster membership (women 60-69yo)

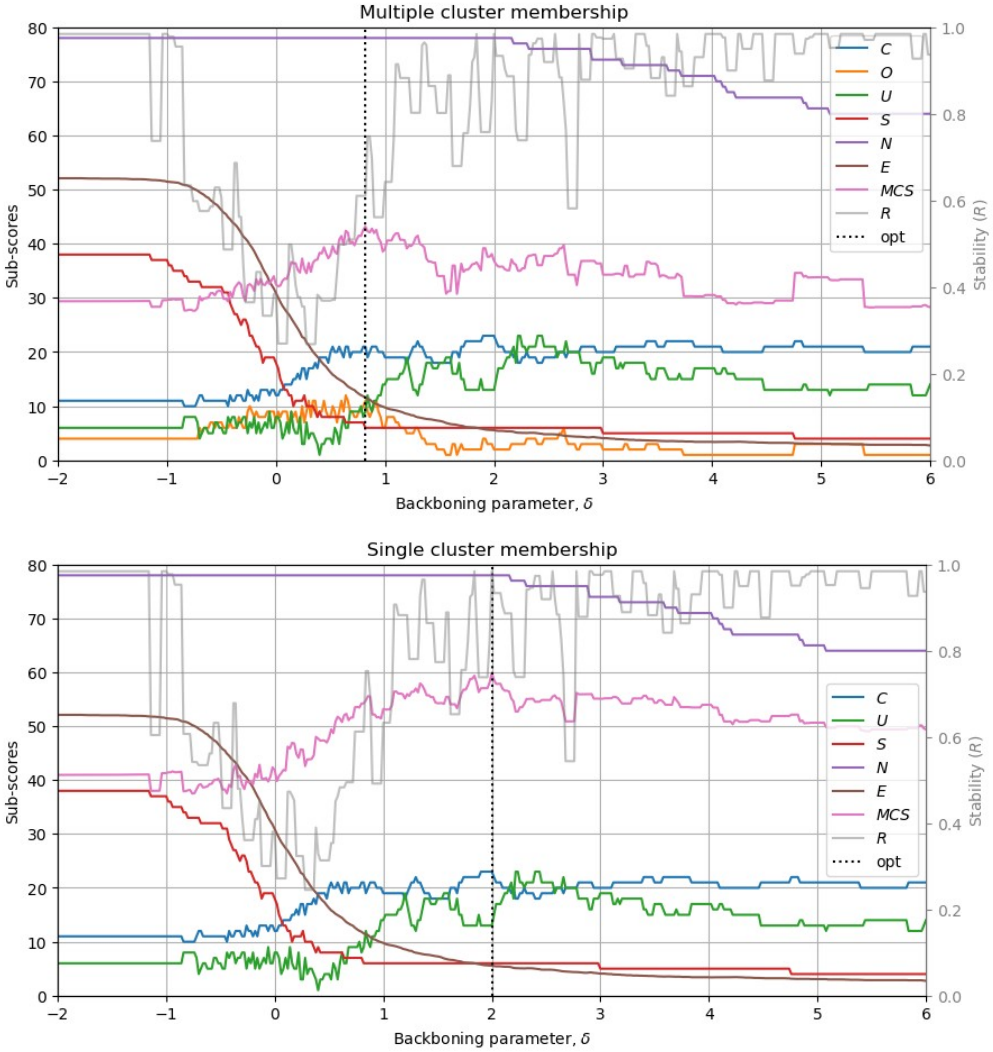


**Figure S4.** Scores of clustering solutions with multiple and single membership clustering algorithms for 60-69 years old women for different values of the backboning parameter δ, in steps of 0.02, and fixed parameters $d_{thresh}=1$ and $cp_{tresh}=0.5$. The lines represent: *C:* the number of clusters in the solution, *O:* the number of conditions with multiple memberships, *U*: the number of conditions not belonging to any cluster, *S:* the size (number of conditions) of the biggest cluster, *N:* The number of nodes in the network, *E:* the average number of connections per node, *MCS:* the *Multimorbidity Clustering* Score, *R:* the stability sub-score, *opt:* the backboning parameter δ that results in the highest *MCS.*

#### S5. Cluster memberships for shown solution comparing multiple to single membership clusters

The clusters for the **multiple-membership** result in Figure 4a are:

- Bipolar Affective Disorder, OCD, Eating Disorders, Alcohol Misuse, PTSD, Substance Misuse
- Heart Valve Disorder, Heart Failure, Atrial Fibrillation, Abdominal Aortic Aneurysm, Cardiac conduction disorder
- Addison's Disease, Connective tissue disease, Inflammatory bowel disease, Psoriasis, Cancer (skin)
- Epilepsy, Multiple Sclerosis, Stroke/TIA, Parkinson's Disease, Dementia
- Glaucoma, Type 1 Diabetes Mellitus, Macular Degeneration, Uveitis, Cataract
- CKD, Polycystic Ovarian Syndrome, Endometriosis, Gout
- Sickle Cell Disease, Thalassaemia, Sarcoidosis, Other haemolytic anaemia
- COPD, Bronchiectasis, Respiratory Failure, Pulmonary Fibrosis
- Polycystic Ovarian Syndrome, Hypertension, Type 2 Diabetes Mellitus, Severely obese
- Other haemolytic anaemia, Cancer (haematological), Aplastic anaemia, Immunodeficiency
- Schizophrenia, Intellectual Disability, OCD, Autism spectrum disorder
- Atrial Fibrillation, Supraventricular Tachycardia, Primary Pulmonary Hypertension, Cardiomyopathy
- Osteoporosis or MOF, Cancer solid organ, Myasthenia Gravis
- Subarachnoid Haemorrhage, Abdominal Aortic Aneurysm, Chronic pancreatitis
- Chronic liver disease, Substance Misuse, HIV
- CHD, Heart Failure, Asbestosis
- COPD, Cystic Fibrosis, Asthma
- Motor Neurone Disease, Epilepsy, Cerebral Palsy
- Hyperparathyroidism, Thyroid Disease, Cancer (skin)
- Diabetic Eye Disease, Peripheral Arterial Disease, Type 1 Diabetes Mellitus
- Anxiety, Cancer (skin), Depression

The conditions belonging to multiple clusters are:

- Cancer (skin): 3
- COPD: 2
- Atrial Fibrillation: 2
- Epilepsy: 2
- Heart Failure: 2
- Substance Misuse: 2
- OCD: 2
- Type 1 Diabetes Mellitus: 2
- Polycystic Ovarian Syndrome: 2
- Other haemolytic anaemia: 2
- Abdominal Aortic Aneurysm: 2

Conditions not belonging to any cluster include Osteoarthritis, Deafness, Diverticular Disease, Neuropathy, Upper GI acid disorder, Venous thromboembolic disease, Rheumatoid Arthritis, Sleep apnoea, Tuberculosis.

The clusters for the **single-membership** result in Figure 4b are:

- Eating Disorders, Substance Misuse, Bipolar Affective Disorder, PTSD, OCD, Alcohol Misuse
- Cardiac conduction disorder, Abdominal Aortic Aneurysm, Heart Failure, Heart Valve Disorder, Atrial Fibrillation
- Addison's Disease, Connective tissue disease, Inflammatory bowel disease, Cancer (skin), Psoriasis
- Dementia, Multiple Sclerosis, Parkinson's Disease, Epilepsy, Stroke/TIA
- Macular Degeneration, Type 1 Diabetes Mellitus, Uveitis, Glaucoma, Cataract
- CKD, Polycystic Ovarian Syndrome, Gout, Endometriosis
- Sickle Cell Disease, Sarcoidosis, Thalassaemia, Other haemolytic anaemia
- COPD, Respiratory Failure, Bronchiectasis, Pulmonary Fibrosis
- Osteoporosis or MOF, Myasthenia Gravis, Cancer solid organ
- Type 2 Diabetes Mellitus, Hypertension, Severely obese
- Cancer (haematological), Immunodeficiency, Aplastic anaemia
- Intellectual Disability, Schizophrenia, Autism spectrum disorder
- Primary Pulmonary Hypertension, Cardiomyopathy, Supraventricular Tachycardia

- Chronic pancreatitis, Subarachnoid Haemorrhage
- Chronic liver disease, HIV
- Asbestosis, CHD
- Asthma, Cystic Fibrosis
- Motor Neurone Disease, Cerebral Palsy
- Hyperparathyroidism, Thyroid Disease
- Peripheral Arterial Disease, Diabetic Eye Disease
- Depression, Anxiety

Conditions not belonging to any cluster are Osteoarthritis, Deafness, Diverticular Disease, Neuropathy, Upper GI acid disorder, Venous thromboembolic disease, Rheumatoid Arthritis, Sleep apnoea, Tuberculosis.

### S6. All clustering sub-scores with open clusters (women 60-69yo)

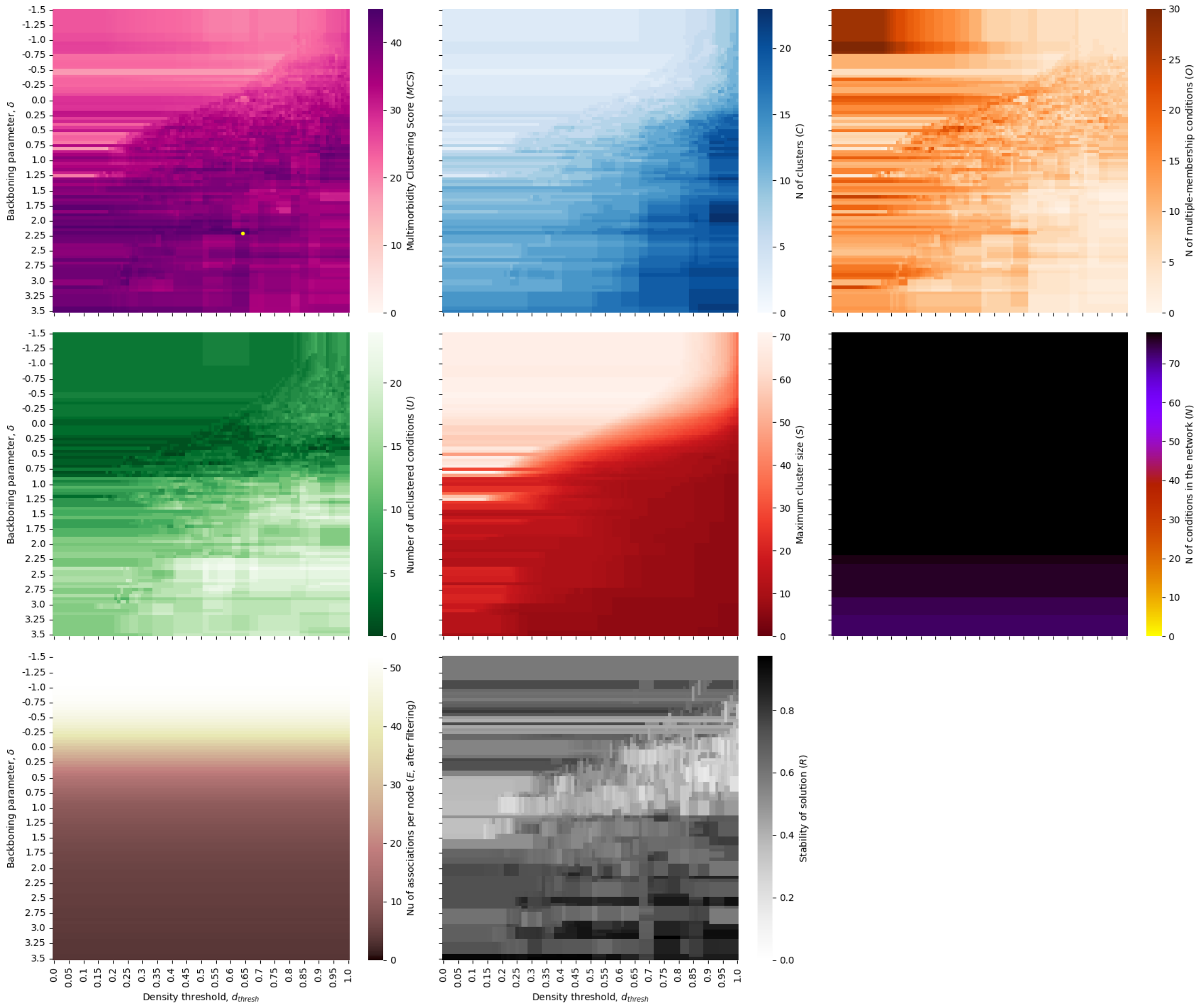


**Figure S6.** Clustering sub-scores for different parameter combinations when clusters are allowed to be ‘open’ (i.e. with nodes not necessarily directly associated to every other node) in women aged 60-69, for a grid of parameter combinations $\delta$-$d_{thresh}$.

#### S7. *MCS* for all age groups with open clusters

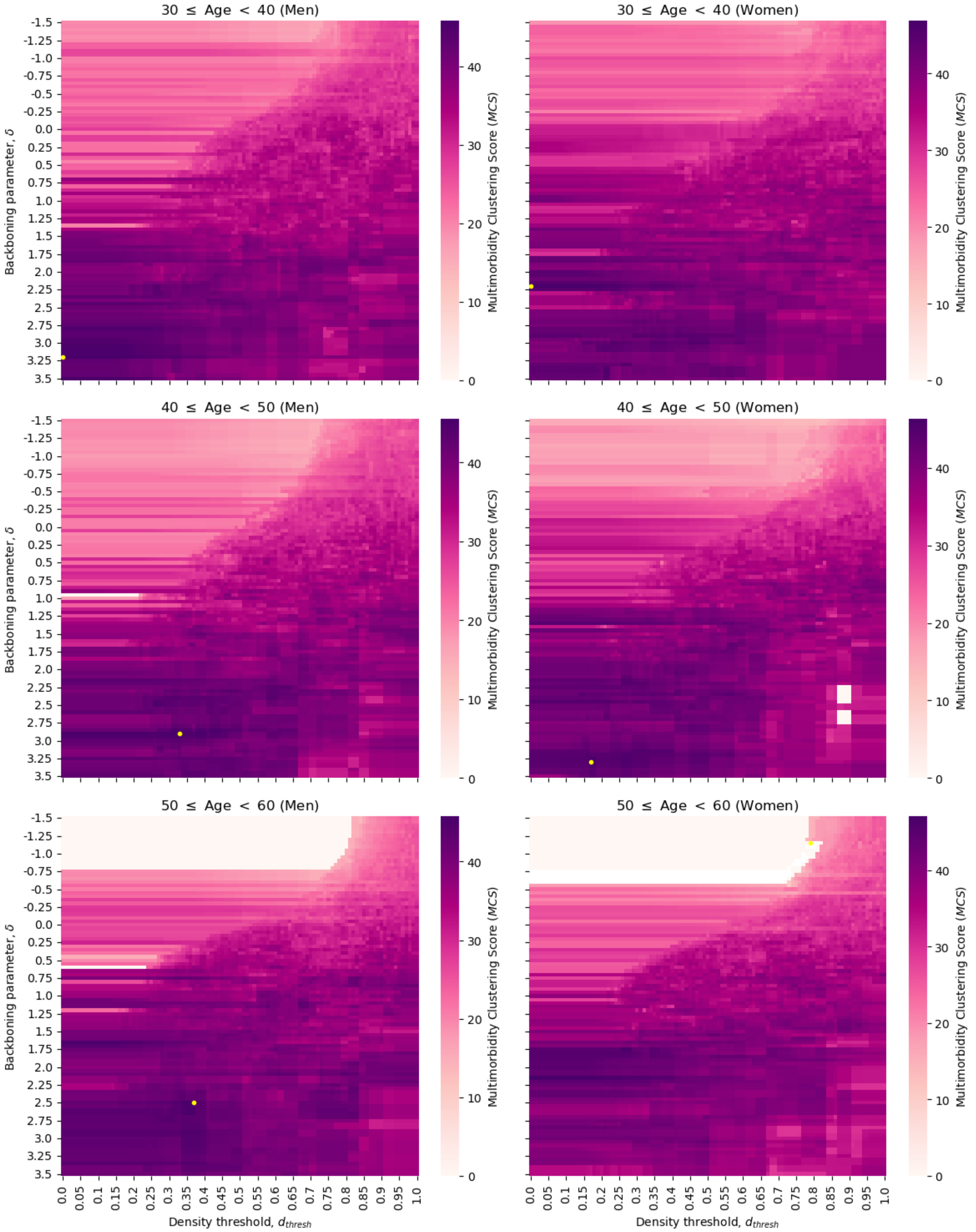

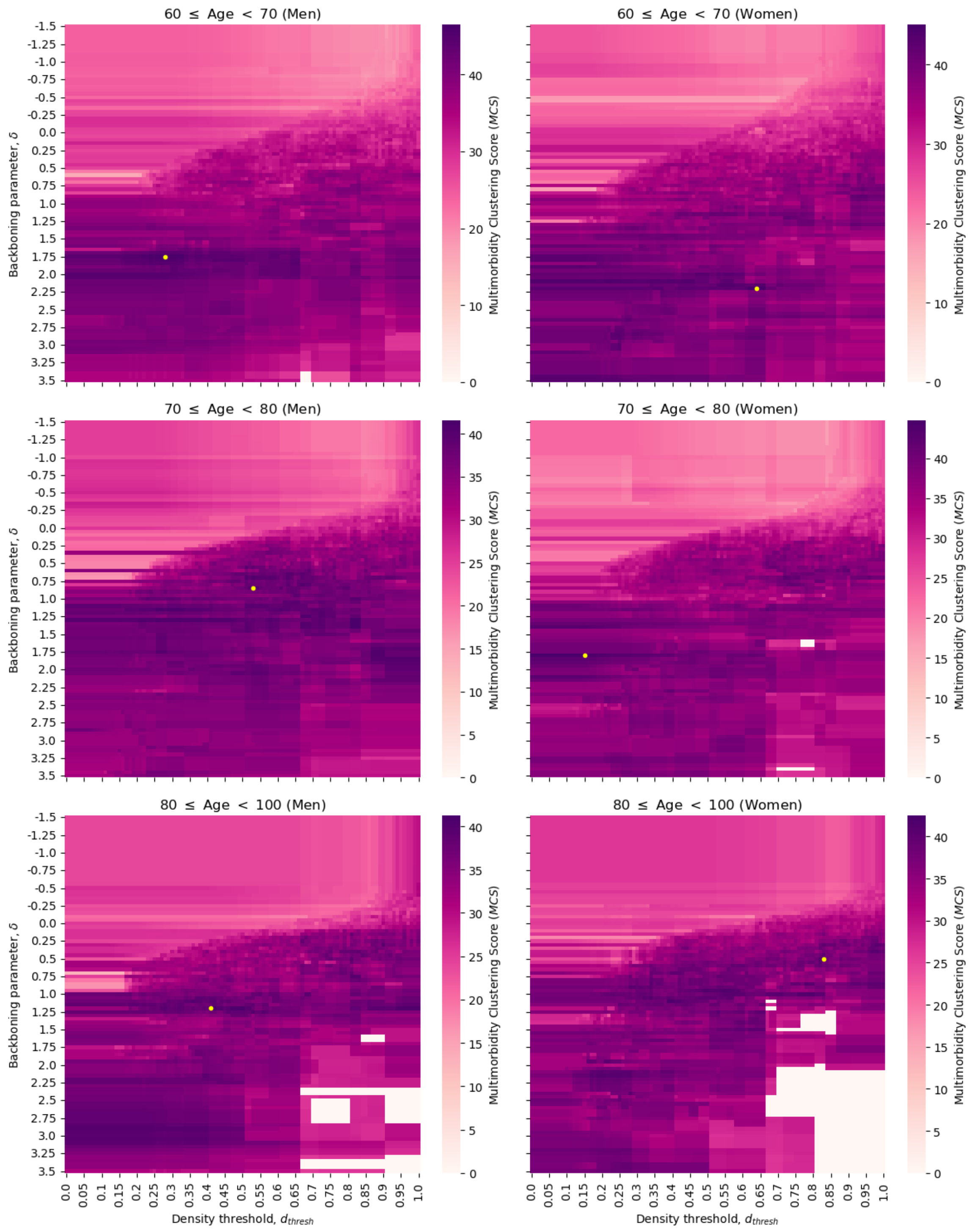


**Figure S7.** *Multimorbidity Clustering Score (MCS)* for different parameter combinations when clusters are allowed to be 'open' (i.e. with nodes not necessarily directly associated to every other node) for all age-sex strata, for a grid of parameter combinations $\delta$-$d_{thresh}$. Yellow dots mark the parameter combination that leads to the highest *MCS* in each age-sex stratum.

### S8. Effect of the cluster-property threshold in open-clusters solutions (women 60-69yo)

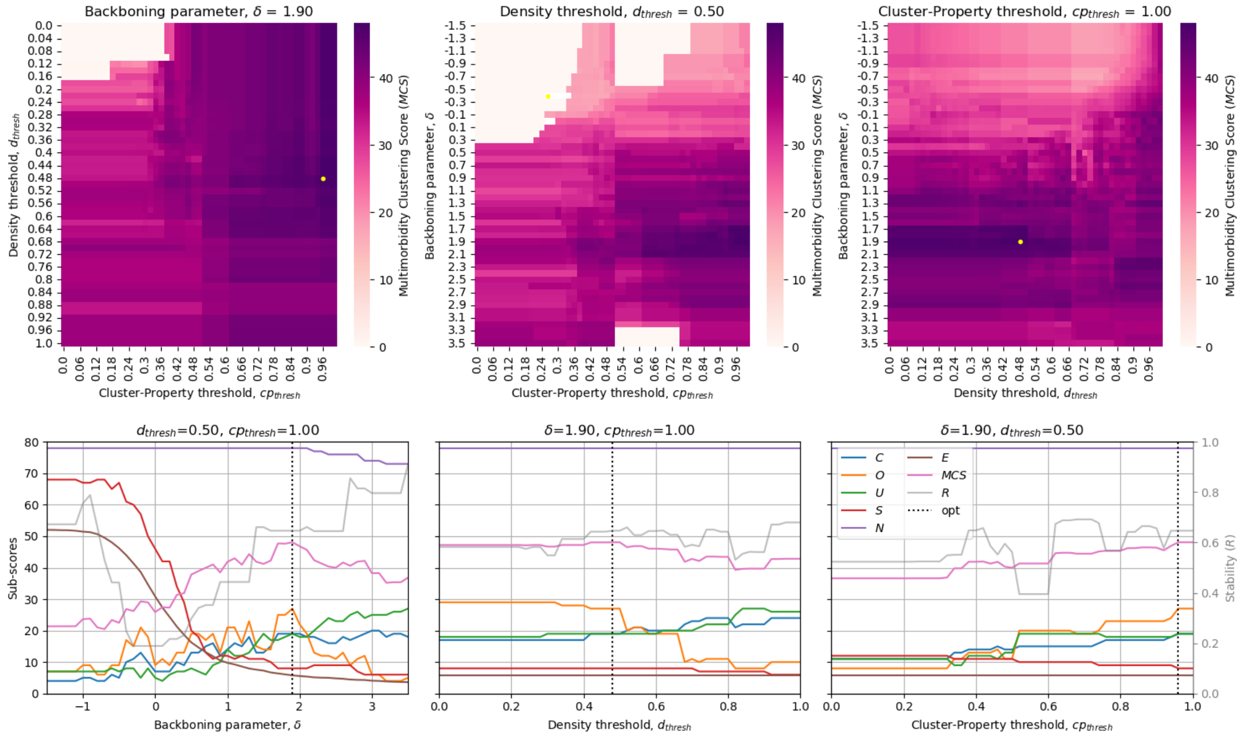


**Figure S8.** Clustering solutions when the cluster-property threshold parameter ($cp_{tresh}$) is allowed to vary and clusters are allowed to be 'open' (i.e. with nodes not necessarily directly associated to every other node), in women aged 60-69. (a) *Multimorbidity Clustering Score (MCS)* for different pairwise parameter combinations while the third parameter is kept fixed at a single value. Yellow dots mark the parameter combination that leads to the highest *MCS*. (b) Sub-scores of clustering solutions for different parameter values of $\delta$, $d_{thresh}$, and $cp_{tresh}$, when the other two parameters are kept fixed at a single value. The lines represent: *C:* the number of clusters in the solution, *O:* the number of conditions with multiple memberships, *U*: the number of conditions not belonging to any cluster, *S:* the size (number of conditions) of the biggest cluster, *N:* The number of nodes in the network, *E:* the average number of connections per node, *MCS:* the *Multimorbidity Clustering* Score*,* *R:* the stability sub-score, *opt:* the parameter that results in the highest *MCS.*

### S9. Different definitions of the stability sub-score with open clusters (women 60-69yo)

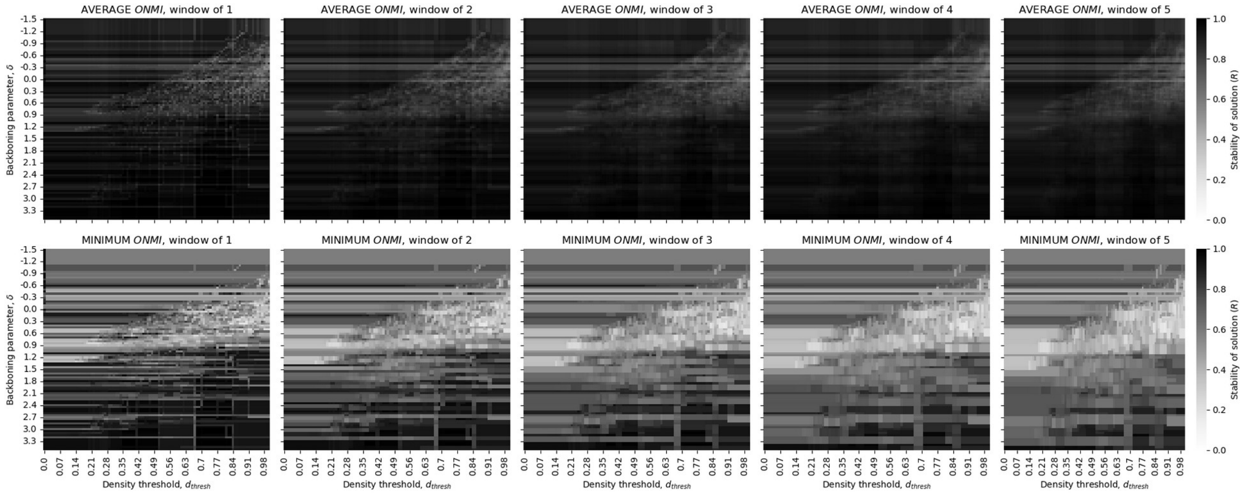


**Figure S9.** Different definitions of the stability sub-score (*R*) of clustering solutions when clusters are allowed to be 'open' (i.e. with nodes not necessarily directly associated to every other node) for 60-69 years old women, for a grid of parameter combinations $\delta$-$d_{thresh}$.

#### S10. Details of CPRD data processing

Data was extracted between June and August 2022 from the CPRD Aurum May 2022 (Version 2022.05.001 of 9/5/2022)[1]. We extracted data for patient identifiers with “acceptable flag” equal to 1 (status “acceptable”) according to CPRD Aurum Denominator Data Specification version 1.2 of 14 January 2022. Linked hospital and death data was extracted in November 2022 from the same CPRD Aurum version (2022.05.001 of 9/5/2022). The CPRD Aurum database May-2022 version contained 41,200,722 research acceptable participants.

In the raw data extraction completed in August 2022 we obtained data for all participants aged 18 years and over on 1st January 2018 who have at least one year’s registration, for a total of 9,131,006 patients.

We excluded patients having sex recorded as I=“3” or U=“4” (hence neither female=”2” nor male=”1”). We excluded patients coming from 29 practices (19 of which were present in our dataset) that CPRD highlighted as duplicated and to be excluded from analyses, as described in CPRD Aurum Data Specification, Version 2.8, Date: 10 August 2022. We excluded patients that had withdrawn their eligibility for linked data as of January 2025.

---

[1]Clinical Practice Research Datalink. (2022). CPRD Aurum May 2022 (Version 2022.05.001). Clinical Practice Research Datalink. https://doi.org/10.48329/t89s-kf12

#### S11. Cluster memberships for shown solution in open regime

The clusters for the **open regime** result in Figure 8 are:

- Dementia, Substance Misuse, Intellectual Disability, Eating Disorders, Autism spectrum disorder, Schizophrenia, Bipolar Affective Disorder, OCD
- Heart Failure, Abdominal Aortic Aneurysm, Primary Pulmonary Hypertension, Cardiac conduction disorder, Atrial Fibrillation, Heart Valve Disorder, Cardiomyopathy
- Sickle Cell Disease, Thalassaemia, Uveitis, Tuberculosis, Sarcoidosis, Other haemolytic anaemia
- Glaucoma, Type 1 Diabetes Mellitus, Uveitis, Macular Degeneration, Cataract
- HIV, Pulmonary Fibrosis, Cystic Fibrosis, Tuberculosis, Bronchiectasis
- Dementia, Motor Neurone Disease, Cerebral Palsy, Autism spectrum disorder, Epilepsy
- HIV, Aplastic anaemia, Immunodeficiency, Cancer (haematological), Other haemolytic anaemia
- Asbestosis, Heart Failure, COPD, Respiratory Failure
- Diabetic Eye Disease, Polycystic Ovarian Syndrome, Thalassaemia, Type 2 Diabetes Mellitus
- Alcohol Misuse, Substance Misuse, Chronic liver disease, Chronic pancreatitis
- Severely obese, Polycystic Ovarian Syndrome, Sleep apnoea, Gout
- Parkinson's Disease, Dementia, Upper GI acid disorder, Multiple Sclerosis
- Anxiety, Eating Disorders, Depression, OCD
- Myasthenia Gravis, Addison's Disease, Cancer solid organ
- PTSD, Endometriosis, Diverticular Disease
- Inflammatory bowel disease, Abdominal Aortic Aneurysm, CHD
- Stroke/TIA, Epilepsy, Subarachnoid Haemorrhage
- Psoriasis, Myasthenia Gravis, Connective tissue disease]']

The conditions belonging to multiple clusters are:

- Dementia: 3
- Epilepsy: 2
- Heart Failure: 2
- Uveitis: 2

- Tuberculosis: 2
- Substance Misuse: 2
- OCD: 2
- Eating Disorders: 2
- Polycystic Ovarian Syndrome: 2
- Thalassaemia: 2
- Other haemolytic anaemia: 2
- Abdominal Aortic Aneurysm: 2
- Myasthenia Gravis: 2
- Autism spectrum disorder: 2
- HIV: 2

Conditions not belonging to any cluster include Hypertension, Osteoarthritis, Asthma, Thyroid Disease, CKD, Osteoporosis or MOF, Neuropathy, Venous thromboembolic disease, Rheumatoid Arthritis, Cancer (skin), Supraventricular Tachycardia, Peripheral Arterial Disease, Hyperparathyroidism.

Deafness was removed from the network as all its associations were fully filtered out after applying the backboning filter.
